# Omics-Based Computational Approaches for Biomarker Identification, Prediction, and Treatment of Long COVID

**DOI:** 10.1101/2025.04.01.25324942

**Authors:** Sindy Pinero, Xiaomei Li, Junpeng Zhang, Marnie Winter, Sang Hong Lee, Thin Nguyen, Lin Liu, Jiuyong Li, Thuc Duy Le

## Abstract

Long COVID, also referred to as post-acute sequelae of COVID-19 (PASC), is a substantial global health concern estimated to have affected over 145 million individuals worldwide. Characterized by persistent and new symptoms extending beyond four weeks from the initial infection—such as fatigue, breathlessness, and cognitive impairments—Long COVID poses significant challenges to healthcare systems due to its chronic nature and diverse clinical manifestations. An urgent need exists to elucidate its underlying mechanisms to facilitate the development of effective diagnostic tools and targeted treatments. This paper provides a comprehensive overview of the current datasets and computational methods to investigate the causes, risk factors, and potential treatments for Long COVID. To understand better the molecular processes driving this condition, we examine various omics data sources, including genomics, epigenomics, transcriptomics, proteomics, and metabolomics datasets. Moreover, we discuss how integrating multi-omics data and using advanced computational techniques—such as machine learning and network analysis—can enhance Long COVID diagnosis, prognosis, and therapeutic strategies. We also emphasize the importance of larger, more diverse cohorts, longitudinal research designs, and cross-cohort validations to strengthen the reliability and applicability of findings. By addressing these crucial aspects, this review aims to advance the development of effective interventions for Long COVID, ultimately improving patient outcomes and alleviating the disease burden.

## 1. Introduction

Long COVID, also known as Post-Acute Sequelae of COVID-19 (PASC), is a significant global health problem, with an estimated 145 million people affected globally during the first two years of the pandemic (Hou *et al*., 2025a; WHO, 2022). The cumulative global incidence of Long COVID is around 400 million individuals, which is estimated to have an annual economic impact of approximately $1 trillion—equivalent to about 1% of the global economy (Al-Aly *et al*., 2024).

The World Health Organization (WHO) and the United States Centers for Disease Control and Prevention (U.S. CDC) define Long COVID based on the persistence of symptoms for at least two months beyond three months post-COVID infection (CDC, 2023; WHO, 2021). In contrast, the United Kingdom’s National Institute for Health and Care Excellence (NICE) describes it as a condition characterized by new and persisting symptoms beyond four weeks (Davis *et al*., 2021; Munblit *et al*., 2022; Petersen *et al*., 2022).

Symptoms like fatigue, breathlessness, cognitive impairment, cardiovascular complications, and autonomic dysfunction affect up to 50% of COVID-19 patients, an infection caused by the Severe Acute Respiratory Syndrome Coronavirus 2 (SARS-CoV-2), with 5% experiencing them for more than three months (CDC, 2023; WHO, 2021). Long COVID is linked to higher risks of chronic diseases, including cardiovascular disorders, diabetes, and neurological issues (Thompson *et al*., 2022). The global burden is substantial, straining healthcare systems and reducing workforce productivity (Balnis *et al*., 2022; Lee *et al*., 2022), with around 71% of patients reporting limitations in daily activities (Durstenfeld *et al*., 2023).

Despite ongoing research, the underlying mechanisms of Long COVID remain uncertain. Various interrelated pathways have been proposed, including persistent viral reservoirs, immune dysfunction, endothelial damage, autoimmunity, and metabolic disruptions (Lai *et al*., 2023; Tsampasian *et al*., 2023). It is not yet clear whether Long COVID follows one or a combination of these mechanisms or if it shares characteristics with other post-viral syndromes. Recent studies have identified significant alterations in immune-related proteins, suggesting persistent inflammation and coagulation abnormalities (Kruger *et al*., 2022; Peppercorn *et al*., 2023; Pretorius *et al*., 2021). Additionally, research has highlighted several risk factors of Long COVID, including age, sex, ethnicity, socioeconomic status, pre-existing conditions (such as asthma, diabetes, and obesity), and the severity of the initial infection (Khullar *et al*., 2023; Lai *et al*., 2023; Tsampasian *et al*., 2023). Mental health conditions, such as depression, have also been linked to increased susceptibility of Long COVID (Selvakumar *et al*., 2023; Su *et al*., 2022a).

Recent advances in computational analysis methods have enabled the processing of large-scale biological data, allowing for a more comprehensive understanding of Long COVID’s complexity through various-omics studies in the pursuit of identifying biomarkers for early diagnosis and targeted treatment (Durstenfeld *et al*., 2023; Munblit *et al*., 2021; O’Keefe *et al*., 2022; Thompson *et al*., 2022; Wu *et al*., 2022). These investigations have included genomic studies, which, through computational analysis, identified the FOXP4 locus (Lammi *et al*., 2023). Additionally, epigenomic studies have used computational approaches to identify 71 differentially methylated regions that persist one year after infection (Balnis *et al*., 2022; Lee *et al*., 2022). Computational analysis of transcriptomic data has revealed 481-38,021 differentially expressed genes (García-Hidalgo *et al*., 2022; Greene *et al*., 2024), while computational processing of proteomic data has identified 162-190 differentially expressed proteins (Peppercorn *et al*., 2023; Woodruff *et al*., 2023).

Previous reviews of Long COVID research have primarily focused on clinical manifestations, biological mechanisms, or specific types of -omics data (Chen *et al*., 2023; Davis *et al*., 2023; Proal *et al*., 2023; Turner *et al*., 2023). While these reviews have provided valuable insights into individual aspects of Long COVID, a comprehensive analysis of computational approaches across all aspects of Long COVID research is currently lacking.

This review focuses on computational approaches for studying Long COVID, analyzing various data sources across different databases. We examine computational methods serving three main purposes: disease characterization, pattern discovery, and therapeutic development. For disease characterization, we analyze how multi-omics data integration and advanced computational techniques enhance our understanding of Long COVID mechanisms (Cervia-Hasler *et al*., 2024; Cheong *et al*., 2023). For pattern discovery and analysis, we examine both supervised methods that predict risk factors with high accuracy (Cervia-Hasler *et al*., 2024; Woodruff *et al*., 2023) and unsupervised approaches like clustering that reveal distinct patient endotypes (An *et al*., 2023). For therapeutic development, we discuss how computational methods aid drug repurposing and treatment optimization (García-Hidalgo *et al*., 2022; Yin *et al*., 2024).

Despite significant advancements in computational analysis, research on Long COVID still faces major challenges that limit the generalizability and interpretability of findings. Small and heterogeneous sample sizes remain a prevalent issue, affecting statistical power and increasing the risk of bias in study outcomes (Su *et al*., 2022a; Thompson *et al*., 2023). The lack of standardized control groups and absence of pre-infection baseline data complicate the identification of reliable biomarkers and disease mechanisms (Cervia-Hasler *et al*., 2024; Yin *et al*., 2024). Additionally, integrating multi-omics datasets presents computational challenges due to batch effects and variations in data processing methods, leading to inconsistencies across studies (Cheong *et al*., 2023; García-Hidalgo *et al*., 2022). Many studies also lack external validation, raising concerns about reproducibility and the clinical applicability of findings (Lammi *et al*., 2023; *et al*., 2023). Addressing these limitations is crucial for developing robust computational models that can enhance our understanding of Long COVID and support the development of targeted diagnostic and therapeutic strategies (Durstenfeld *et al*., 2023; Munblit *et al*., 2021).

## 2. Background

Long COVID is a complex, multisystem condition characterized by persistent symptoms following acute COVID-19 infection. Studies have demonstrated that post-COVID-19 individuals face increased risks across multiple organ systems: cardiovascular disease, diabetes, and circulatory impairment, along with abnormal ventilatory patterns (Mancini *et al*., 2021; Xie *et al*., 2022; Xie & Al-Aly, 2022). A significant subset of these patients fulfills the 2003 Canadian Consensus Criteria for Myalgic encephalomyelitis/chronic fatigue syndrome (ME/CFS), exhibiting disease severity and symptom burden comparable to non-COVID-19 ME/CFS patients (Kedor *et al*., 2022). Autonomic dysfunction is also highly prevalent in the Long COVID population, independent of the severity of acute COVID-19 illness (Larsen *et al*., 2022). Symptoms of Long COVID can last for years, with some conditions expected to be lifelong (Cairns & Hotopf, 2005; Demko *et al*., 2022), and multiple patients experience reduced work capacity due to continued illness (Munblit *et al*., 2021).

Multiple overlapping mechanisms have been proposed to explain the pathophysiology of Long COVID, including persistent viral reservoirs, immune dysregulation, autoimmunity, reactivation of latent viral infections, and alterations in the gut microbiome (Arthur *et al*., 2021; Buonsenso *et al*., 2022; Glynne *et al*., 2022; Klein *et al*., 2023; Liu *et al*., 2022; Manoharan & Ying, 2023; Peluso *et al*., 2021; Phetsouphanh *et al*., 2022; Proal *et al*., 2023; Proal & VanElzakker, 2021; Shikova *et al*., 2020; Su *et al*., 2022a; Swank *et al*., 2023; Zubchenko *et al*., 2022). The persistence of the virus or its components is hypothesized to activate a dysregulated immune system with increased release of pro-inflammatory cytokines, leading to chronic low-grade inflammation and multiorgan symptomatology (Buonsenso *et al*., 2022; Proal *et al*., 2023). Multiple forms of immune dysregulation have been identified as key features of Long COVID, including T cell alterations, elevated programmed cell death protein 1 (*PD-1*) expression, activated innate immune cells, distinct cytokine patterns, and the development of autoantibodies against various receptors (Arthur *et al*., 2021; Glynne *et al*., 2022; Klein *et al*., 2023; Peluso *et al*., 2021; Phetsouphanh *et al*., 2022; Proal & VanElzakker, 2021; Su *et al*., 2022a).

Vascular and organ damage have been reported as key manifestations of Long COVID, including endothelial dysfunction, thrombosis, and organ-specific damage, and long-term effects on cardiovascular, renal, and other systems (Bowe *et al*., 2021; Charfeddine, 2021; Dennis *et al*., 2023; Osiaevi *et al*., 2023; Patel *et al*., 2022; Pretorius *et al*., 2021; Puntmann *et al*., 2020; Raman *et al*., 2022; Turner *et al*., 2023; Xie *et al*., 2022). Persistent capillary rarefaction, endothelial dysfunction, and the presence of large anomalous deposits resistant to fibrinolysis in plasma samples have been observed in patients with Long COVID (Charfeddine, 2021; Osiaevi *et al*., 2023; Pretorius *et al*., 2021; Turner *et al*., 2023).

Neurological and cognitive manifestations, including widespread neurocognitive deficits, brain structural changes, and neuropathologies, have been widely reported in patients with Long COVID (Apple *et al*., 2022; Ceban *et al*., 2022; Cysique *et al*., 2023; Davis *et al*., 2021; Guedj *et al*., 2021; Holdsworth *et al*., 2022; Hugon *et al*., 2022; Monje & Iwasaki, 2022; Reiken *et al*., 2022; Taquet *et al*., 2022). Meta-analyses have revealed the persistent experience of fatigue and cognitive dysfunction following the acute COVID-19 period (Ceban *et al*., 2022; Davis *et al*., 2021). Objective cognitive impairment has been associated with reduced work capacity, anosmia, abnormalities in the kynurenine pathway, and the activation of neuropathological pathways typically associated with Alzheimer’s disease (Cysique *et al*., 2023; Holdsworth *et al*., 2022; Reiken *et al*., 2022). Brain Positron Emission Tomography (PET) studies have demonstrated hypometabolism in various brain regions, with these abnormalities being associated with patients’ symptoms (Guedj *et al*., 2021; Hugon *et al*., 2022). The underlying pathophysiological processes that may contribute to this emerging neurological health crisis include viral factors, host factors, and downstream impacts (Monje & Iwasaki, 2022).

Beyond neurological manifestations, emerging evidence suggests that the long-term sequelae of SARS-CoV-2 infection may extend to oncological implications, potentially increasing the risk of cancer development and progression. The potential mechanisms linking Long COVID and cancer include chronic inflammation, immune dysregulation, activation of oncogenic pathways, tissue damage, cellular senescence, and alterations in the gut microbiome (Amiama-Roig *et al*., 2023; Conti *et al*., 2023; Costanzo *et al*., 2023; Jahankhani *et al*., 2023; Rahimmanesh *et al*., 2022; Saini & Aneja, 2021). The persistent pro-inflammatory state, characterized by elevated levels of cytokines such as interleukin-1 beta (IL-1*β*), interleukin-6 (IL-6), interleukin-13 (IL-13), interleukin-17A (IL-17A), tumor necrosis factor-alpha (TNF*α*), interferon gamma-induced protein-10 (IP-10), and granulocyte colony-stimulating factor (G-CSF), may create a microenvironment conducive to cancer development (Conti *et al*., 2023). T cell exhaustion, marked by the overexpression of *PD-1* and programmed death-ligand 1 (PD-L1), has been observed in patients with Long COVID and may facilitate tumor immune escape (Conti *et al*., 2023). SARS-CoV-2 has been shown to modulate oncogenic pathways, such as the mammalian target of rapamycin (mTOR) pathway and p38 mitogen-activated protein kinase (MAPK) pathway, and interact with tumor suppressor proteins like tumor protein p53 (p53) and breast cancer type 1/2 susceptibility protein (BRCA1/2), potentially contributing to genomic instability and aberrant cell growth (Conti *et al*., 2023). The persistence of viral antigens and RNA in various tissues, even months after the acute infection, may lead to chronic low-grade inflammation and tissue damage, creating a favorable environment for cancer development (Amiama-Roig *et al*., 2023). Cellular senescence, characterized by the senescence-associated secretory phenotype (SASP) and the accumulation of senescent cells, has been linked to both Long COVID and cancer, with the chronic inflammation induced by SASP potentially contributing to tumorigenesis (Amiama-Roig *et al*., 2023). Alterations in the gut microbiome composition, such as the depletion of beneficial bacteria and the proliferation of opportunistic pathogens, have been observed in patients with Long COVID and may influence cancer susceptibility and prognosis (Conti *et al*., 2023).

The identified biological mechanisms underlying Long COVID represent significant advances in our understanding of this condition. However, the precise etiology remains under investigation, with key research priorities including identifying reliable biomarkers, clarifying risk factors, and developing targeted therapeutic approaches. Furthermore, Long COVID presents a spectrum of manifestations with multiple contributing factors varying among individuals. Genetic studies, mainly when analyzed through advanced computational methods, may provide crucial insights into these biological mechanisms, potentially leading to more targeted treatments and preventative measures.

Given the complexity and heterogeneity of Long COVID, computational methods have become increasingly essential for analyzing large-scale datasets, identifying patterns, and generating new hypotheses. As noted earlier, while several reviews have focused on clinical aspects and biological mechanisms of Long COVID, a comprehensive analysis of the generated omics datasets and computational approaches in Long COVID research is currently lacking. This review aims to bridge this gap by systematically cataloging available datasets and examining how different computational methods are being applied across the full spectrum of Long COVID research, from mechanistic understanding to therapeutic development. By providing a consolidated resource of current datasets and their analytical frameworks, we aim to facilitate more efficient data discovery and promote more effective use of computational approaches to accelerate progress in understanding and treating this condition. We begin by examining available data sources and their computational analysis, followed by a detailed discussion of specific computational approaches and their applications in extracting meaningful insights from these complex datasets.

## 3. Computational Approaches and Omics Data in Long COVID Research

Understanding Long COVID at a molecular level requires advanced computational approaches to analyze high-dimensional biological data, identify biomarkers, and develop predictive models. This review aims to systematically assess the current omics datasets and computational methods used in Long COVID research, focusing on how bioinformatics, statistical modeling, and machine learning (ML) techniques have been applied to study the disease. By evaluating existing methodologies, we seek to identify key computational strategies, assess their impact, and highlight methodological gaps that require further exploration.

We define “computational methods” as algorithmic approaches, statistical techniques, and software tools used to analyze, interpret, and derive insights from biological data. These include data processing pipelines, statistical analysis frameworks, machine learning algorithms, network analysis tools, and specialized bioinformatics software for omics data analysis. In our review, we categorize these methods by analytical tasks (such as primary analysis, dimensionality reduction, and pathway analysis) to structure the diverse computational landscape applied to Long COVID research.

To comprehensively capture relevant studies, we searched multiple scientific databases (Google Scholar, PubMed) and biological data repositories (Gene Expression Omnibus, European Nucleotide Archive, Sequence Read Archive, and COVID-19 Data Portal).

We conducted a systematic review across multiple databases (Google Scholar, PubMed, Gene Expression Omnibus, European Nucleotide Archive, Sequence Read Archive, and COVID-19 Data Portal) analyzing studies up to March 3, 2025 (for complete search methodology, links and terms, see Supplementary Materials 1 and 2 (SM1 and SM2)). Our analysis involved both generating new molecular datasets and computational analysis of existing data across various molecular data types, including gene expression (microarray, bulk, and single-cell RNA sequencing), genome-wide association studies (GWAS), epigenome-wide association studies (EWAS), proteomics, metabolomics, and multi-omics. This combined approach of data generation and computational analysis allowed for comprehensive investigation of Long COVID molecular mechanisms. These studies utilized differential expression analysis, pathway enrichment, network-based modeling, ML classification, integrative multi-omics approaches, and statistical frameworks to uncover molecular mechanisms underlying Long COVID.

Computational methods are essential in understanding disease mechanisms, improving biomarker discovery, and developing predictive models for Long COVID prognosis and treatment response. This review is a comprehensive resource on existing methodologies, guiding future research toward more effective integration of computational and multi-omics approaches to enhance our understanding of Long COVID’s molecular basis.

### 3.1. Study Characteristics and Omics Data Distribution

Our systematic review highlights distinct trends in using computational approaches across various omics studies in Long COVID research. Genomics studies, particularly genome-wide association studies (GWAS), involve the largest cohorts, reflecting the feasibility of large-scale genetic analyses. Transcriptomics studies (including bulk and single-cell RNA sequencing) tend to have smaller sample sizes but demonstrate the highest diversity in sample types, ranging from lung tissue to blood components. Proteomics and metabolomics studies often analyze medium-to-large cohorts and predominantly utilize blood samples, with proteomics showing structured temporal sampling patterns. Epigenomics studies (EWAS) remain less common but provide insights into regulatory modifications associated with Long COVID. Multi-omics approaches, which integrate multiple data types, exhibit the highest variability in cohort sizes and follow-up durations, emphasizing their potential for a comprehensive understanding of Long COVID pathophysiology.

Across all omics studies, peripheral blood emerges as the most frequently used sample type, likely due to its accessibility and relevance for systemic analyses. Longitudinal studies, particularly in proteomics, transcriptomics, and multi-omics, tend to include larger cohorts and extended follow-up periods, allowing for a more detailed characterization of Long COVID’s molecular alterations over time.

A genomics study by Lammi *et al*. (2023) employed a large-scale GWAS approach analyzing 6,450 Long COVID cases, 46,208 COVID controls, and 1,093,955 non-COVID controls, representing the largest cohort in our review. Similarly, Chaudhary *et al*. (2024) conducted a GWAS study analyzing 53,764 Long COVID cases and 120,688 COVID controls, identifying three genome-wide significant loci associated with Long COVID.

Figure 1 illustrates the distribution of sample sizes across transcriptomics, proteomics, metabolomics, and multi-omics studies in Long COVID research. Genomics studies were not included in this figure, as their sample sizes were substantially larger than other omics datasets, making direct visual comparisons less informative. Multi-omics and proteomics studies typically involve larger cohorts when compared with transcriptomics. The sample size distribution (Fig. 1B) reveals substantial variation across omics types, with multi-omics studies showing the highest variability and several outliers, particularly in Long COVID cases.

**Fig. 1.**
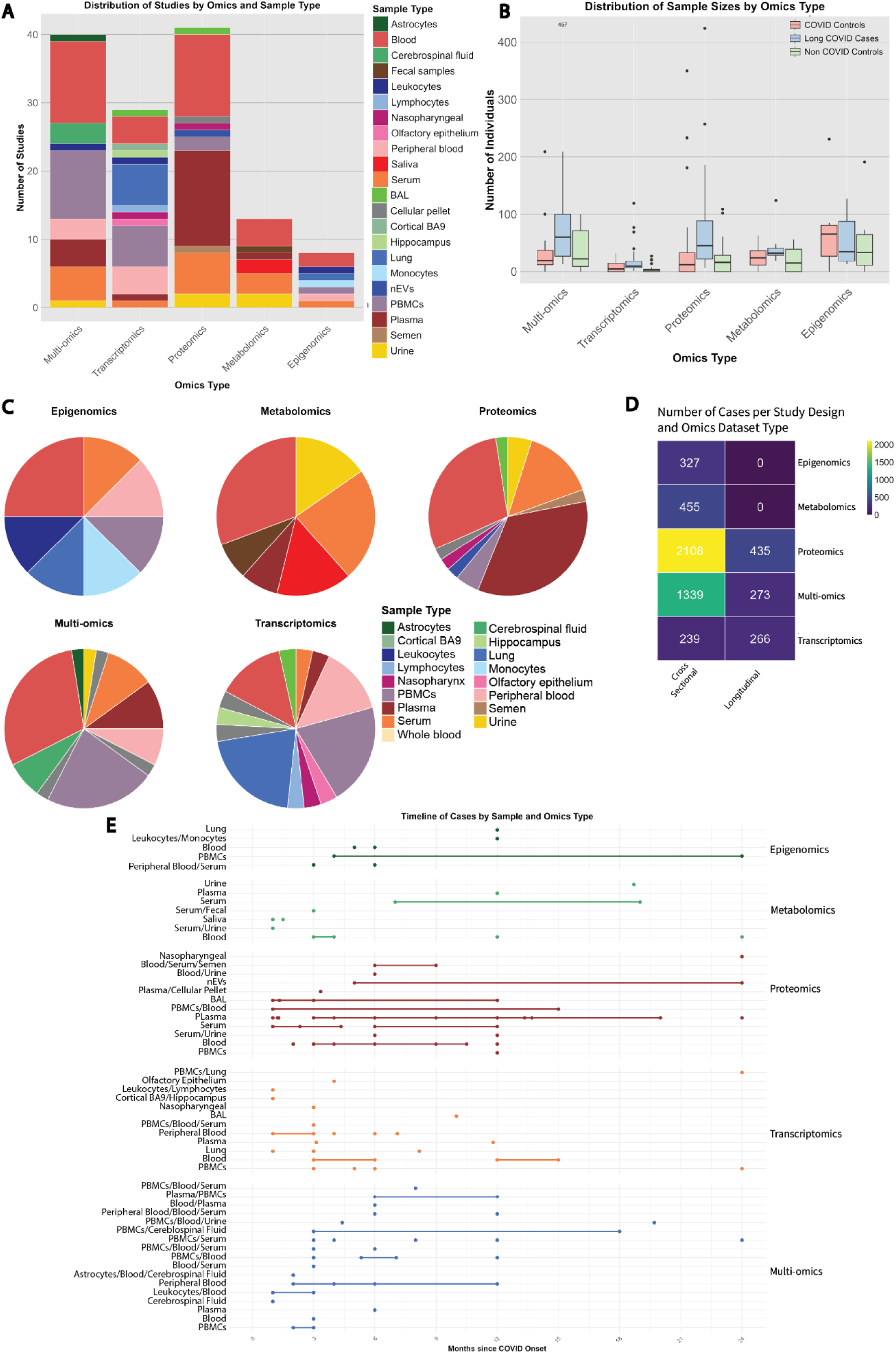
Overview of multi-omics studies in Long COVID. (A) Distribution of studies by omics type and sample source (B) Distribution of sample sizes across omics studies stratified by COVID status (C) Proportion of sample types used in the omics studies (D) Number of cases analyzed in cross-sectional and longitudinal studies for each omics type (E) Timeline of cases based on sample source and omics type in months since COVID onset Abbreviations: BAG: Brain Associated Glycoprotein (in Cortical BAG); BAL: Bronchoalveolar Lavage; COVID: Coronavirus Disease; nEVs: Neuronal Extracellular Vesicles; Multi-omics: Multiple omics data integration; PBMCs: Peripheral Blood Mononuclear Cells

Multi-omics studies show the highest median number of Long COVID cases (1339 in cross-sectional studies, 273 in longitudinal studies), followed by proteomics (2108 in cross-sectional, 435 in longitudinal) and metabolomics (455 in cross-sectional) as shown in Fig. 1D. This observation aligns with the growing emphasis on integrated approaches in Long COVID research, as multi-omics studies often require diverse datasets and comprehensive analysis pipelines. Larger sample sizes in these studies may result from the need for statistical power when integrating multiple molecular layers, enabling more robust conclusions. The proportional analysis (Fig. 1C) demonstrates distinct patterns in sample type utilization across different omics platforms, with notable differences in the distribution of tissue and blood-derived samples. Sample types vary across approaches, with transcriptomics showing the greatest diversity from lung tissue to blood components. Peripheral blood is the most predominant sample type across all methodologies (Fig. 1A), likely due to its accessibility and potential to reflect systemic changes. The analysis reveals a particular emphasis on blood-derived components (PBMCs, plasma, serum) in multi-omics studies, suggesting these samples’ utility in capturing multiple molecular dimensions of Long COVID pathophysiology.

The timeline analysis reveals that most studies collected samples within the first 24 months after COVID onset, with some extending beyond this period. From a computational perspective, the variability in sample collection time points introduces challenges in integrating and comparing datasets, particularly in time-series analyses. Figure 1E presents the distribution of studies based on the number of months post-infection, with each point representing a distinct sampling timepoint and horizontal lines indicating the total study duration. The maximum follow-up periods observed reach approximately 36-40 months in some multi-omics and proteomics studies, providing valuable long-term molecular data.

Longitudinal analyses are prominent in proteomics, transcriptomics, and multi-omics research, primarily utilizing blood samples. The temporal distribution shows varying study durations across sample types, with multi-omics studies demonstrating the most extended follow-up periods, some reaching beyond 36 months. Furthermore, some proteomics studies exhibit consistent sampling intervals, particularly in PBMC and plasma analyses, suggesting structured temporal monitoring of protein-level changes. Our analysis reveals that longer study durations correlate with increased Long COVID case numbers, suggesting the importance of extended follow-up periods to fully characterize Long COVID manifestations. The cross-platform comparison (Fig. 1D) indicates a substantial disparity in case numbers between cross-sectional and longitudinal studies, with longitudinal designs generally incorporating larger cohorts for comprehensive temporal analysis, as seen in proteomics (2108 vs. 435 cases) and multi-omics (1339 vs. 273 cases) approaches.

For additional details of each study, including specific assay methodologies and data pre-process methods, refer to Table S1 in the SM2. This resource will be continuously updated as new studies emerge in this rapidly evolving field.

### 3.2. Long COVID Biomarkers

Figure 2 and Table 1 provide complementary perspectives on multi-level omics approaches in Long COVID research. While the table details specific computational methods and quantitative findings across various omics domains, the figure visually illustrates how these approaches interconnect, from genomic variants (FOXP4) to epigenetic modifications (71 DMRs in promoters), transcriptomic changes (immune dysregulation), proteomic alterations (complement/coagulation activation), and metabolic disruptions (mitochondrial dysfunction). Together, they demonstrate how integrated multi-omics data enable patient classification into distinct endotypes and inform potential therapeutic strategies targeting specific molecular pathways. This comprehensive molecular characterization connects biological signatures to the diverse clinical manifestations of Long COVID—including cognitive, respiratory, and autonomic symptoms—providing a foundation for precision medicine approaches to diagnosis and treatment.

**Fig. 2.**
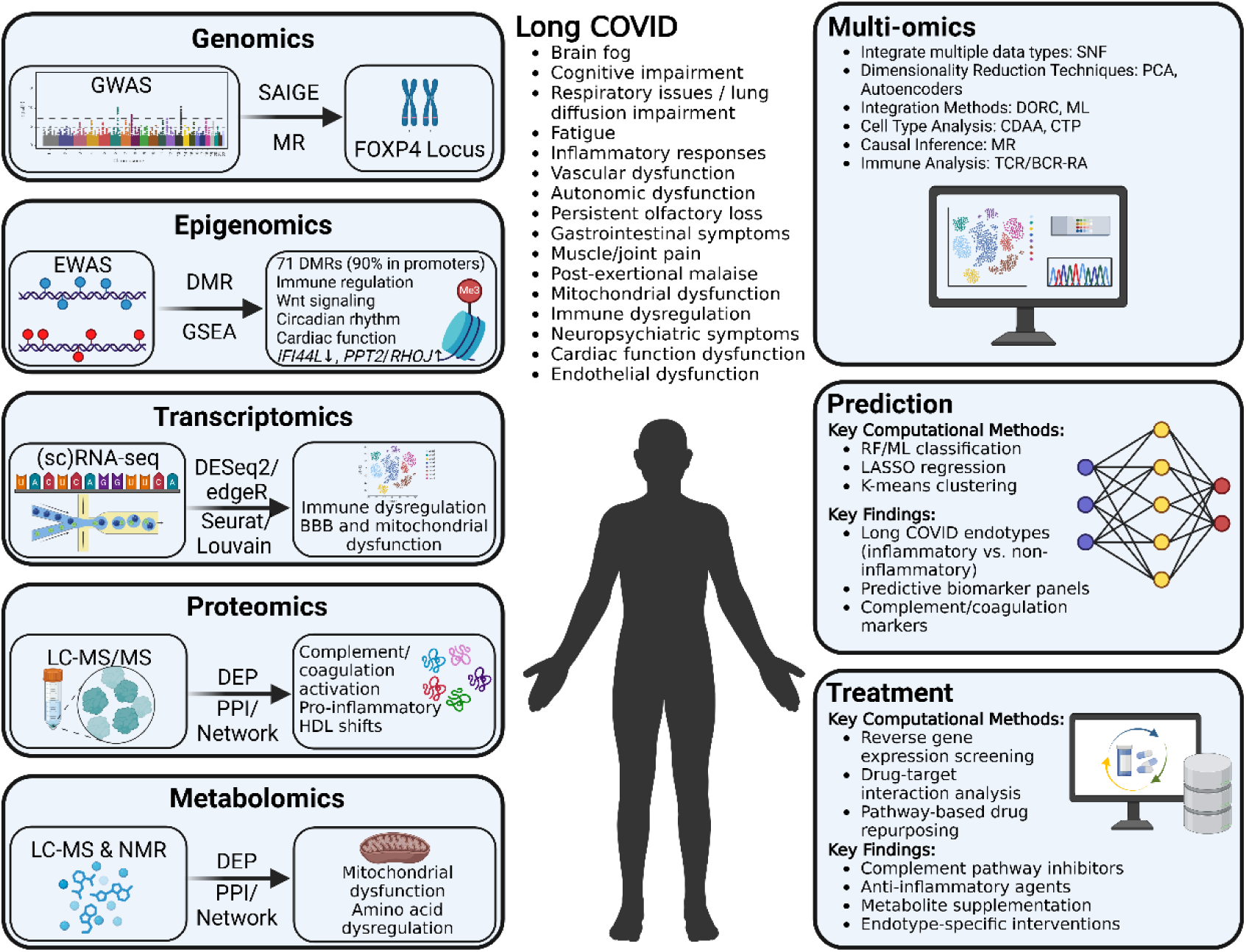
Multi-Level Omics Approaches in Long COVID Research: Computational Methods and Key Findings. Integration of diverse omics approaches for understanding Long COVID mechanisms and developing targeted interventions Different panels show the computational workflows and key findings from genomics, epigenomics, transcriptomics, proteomics, metabolomics, and multi-omics integration along with prediction models and treatment discovery approaches Abbreviations: GWAS: Genome-Wide Association Study; SAIGE: Scalable and Accurate Implementation of GEneralized mixed model; MR: Mendelian Randomization; EWAS: Epigenome-Wide Association Study; DMR: Differentially Methylated Region; GSEA: Gene Set Enrichment Analysis; scRNA-seq: single-cell RNA sequencing; DESeq2: Differential Expression Sequencing 2; BBB: Blood-Brain Barrier; LC-MS/MS: Liquid Chromatography-Mass Spectrometry/Mass Spectrometry; DEP: Differentially Expressed Proteins; PPI: Protein-Protein Interaction; HDL: High-Density Lipoprotein; NMR: Nuclear Magnetic Resonance; SNF: Similarity Network Fusion; PCA: Principal Component Analysis; DORC: Detection of Regulatory Chromatin; CDAA: Cell-type Differential Abundance Analysis; CTP: Cell Type Profiling; TCR/BCR-RA: T-Cell Receptor/B-Cell Receptor Repertoire Analysis; RF/ML: Random Forest/Machine Learning; LASSO: Least Absolute Shrinkage and Selection Operator Created in BioRender (https://BioRender.com/c92d367)

**Table 1.**
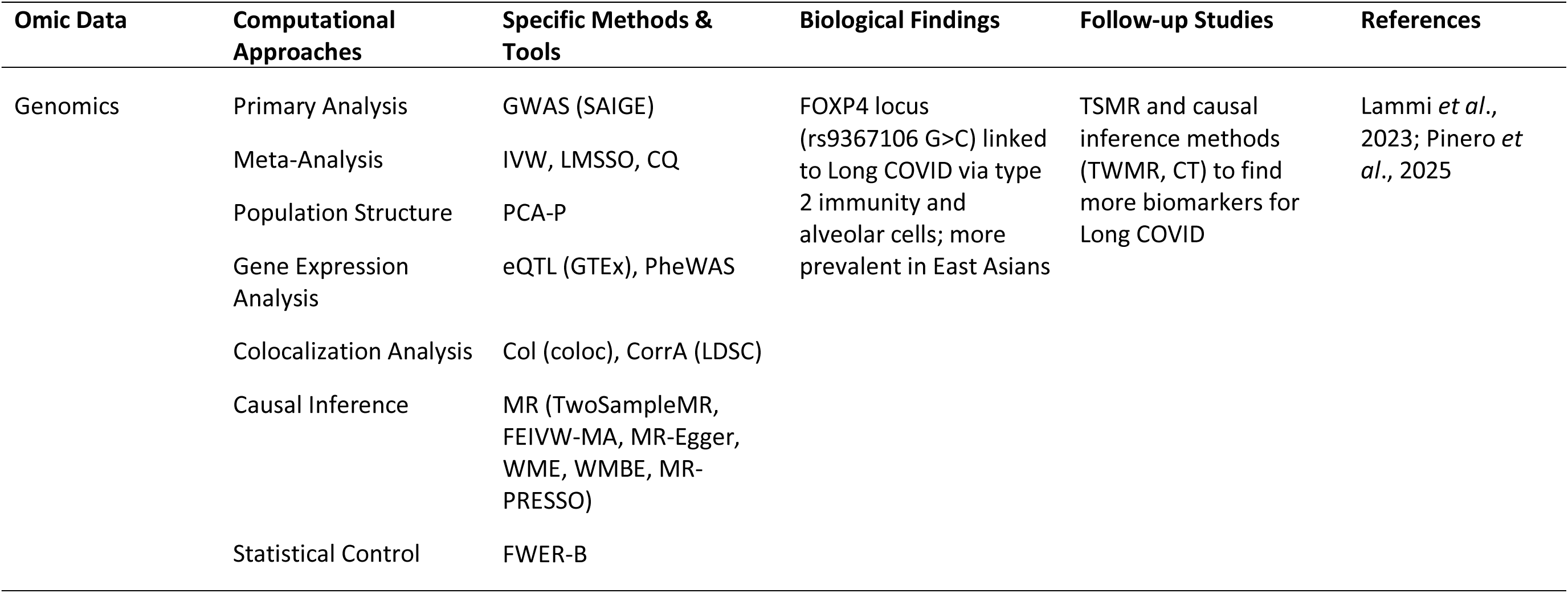
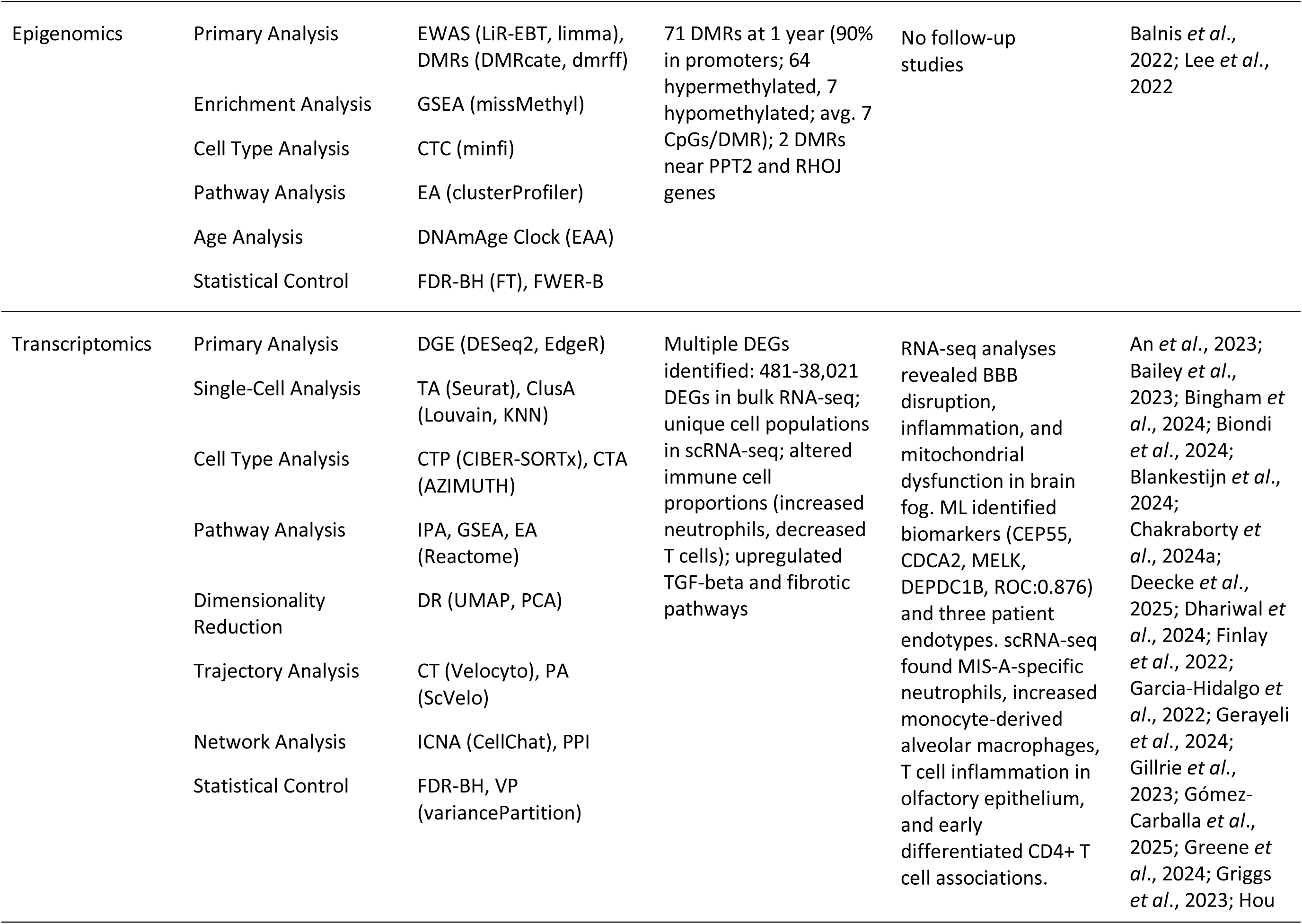
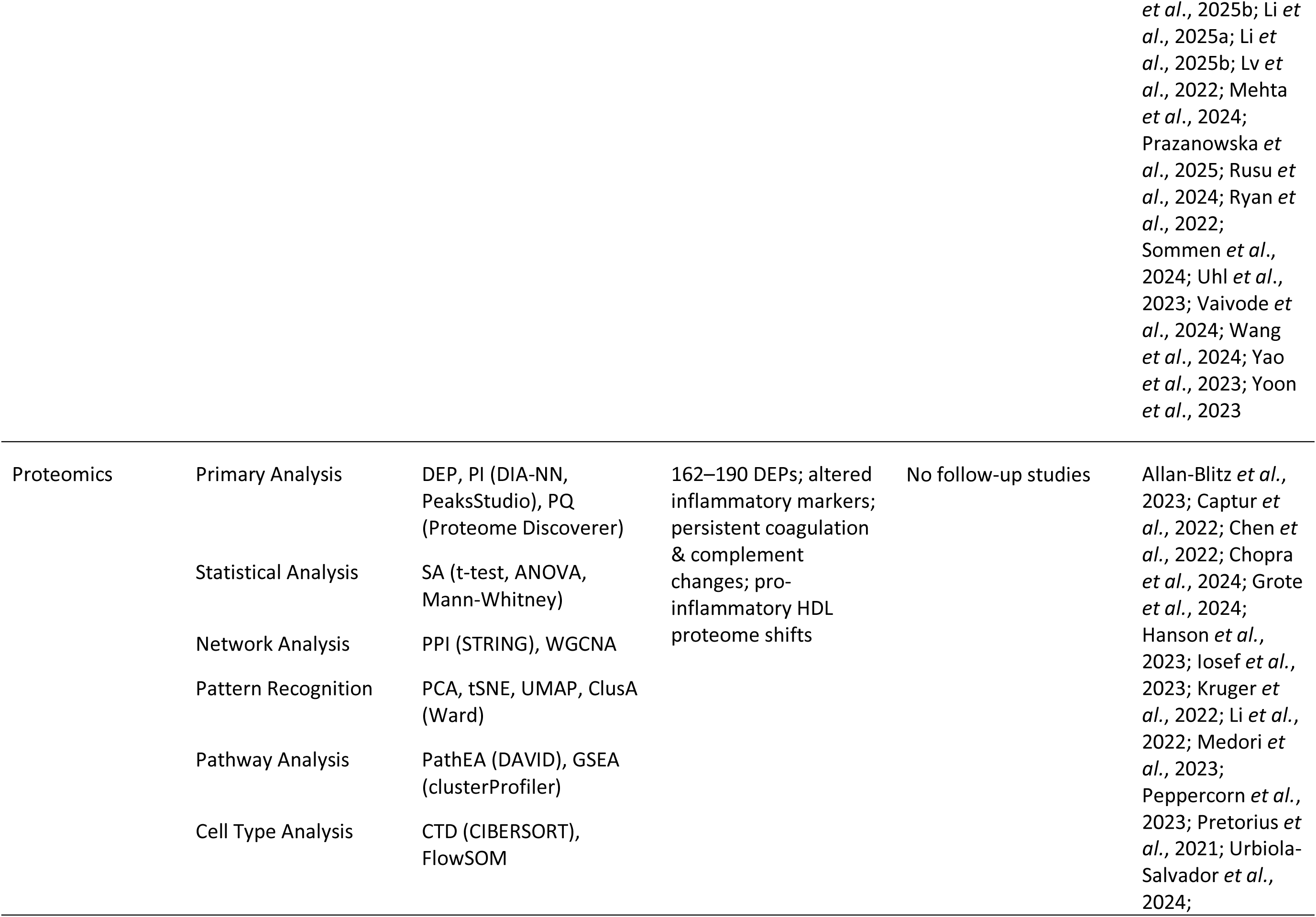
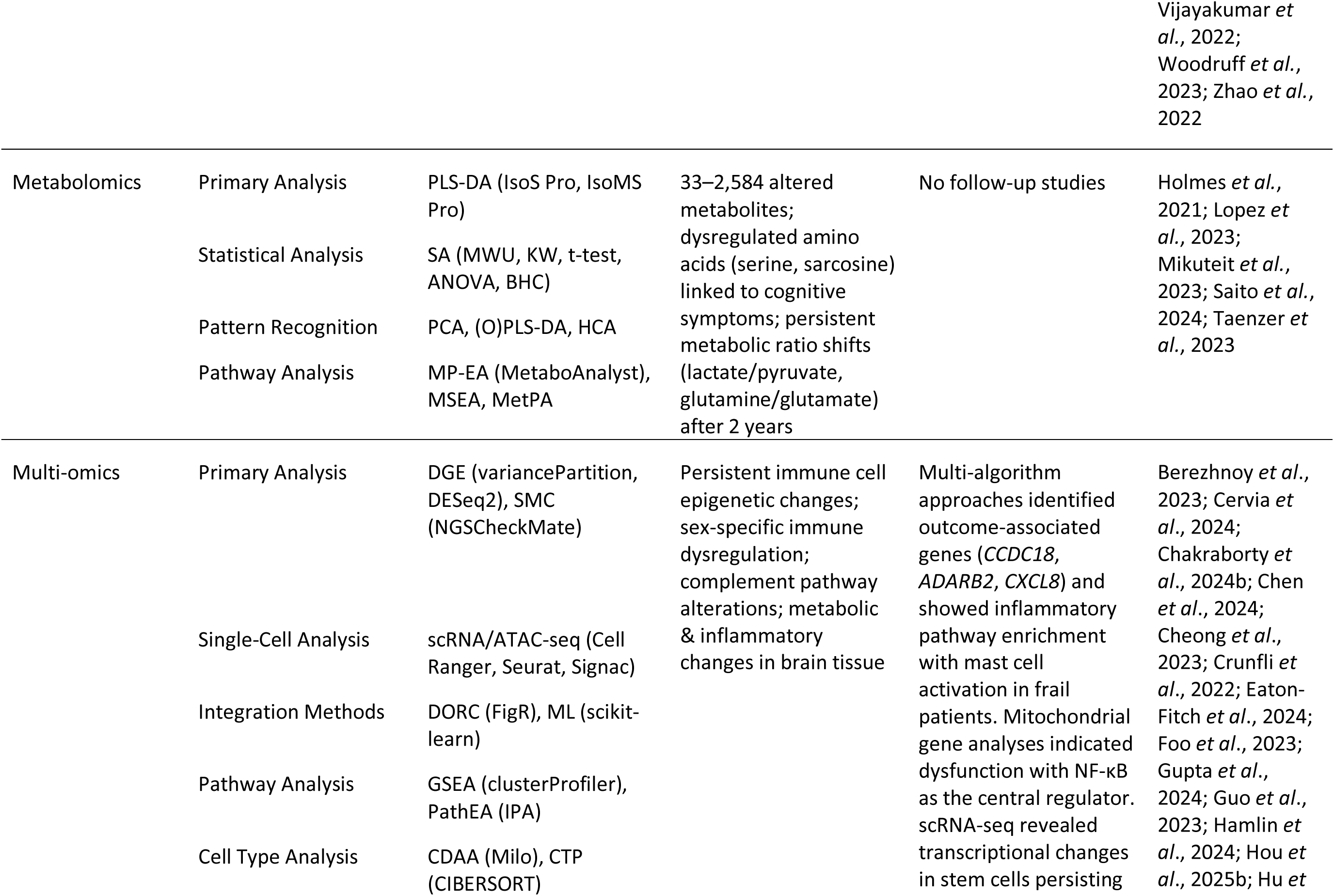
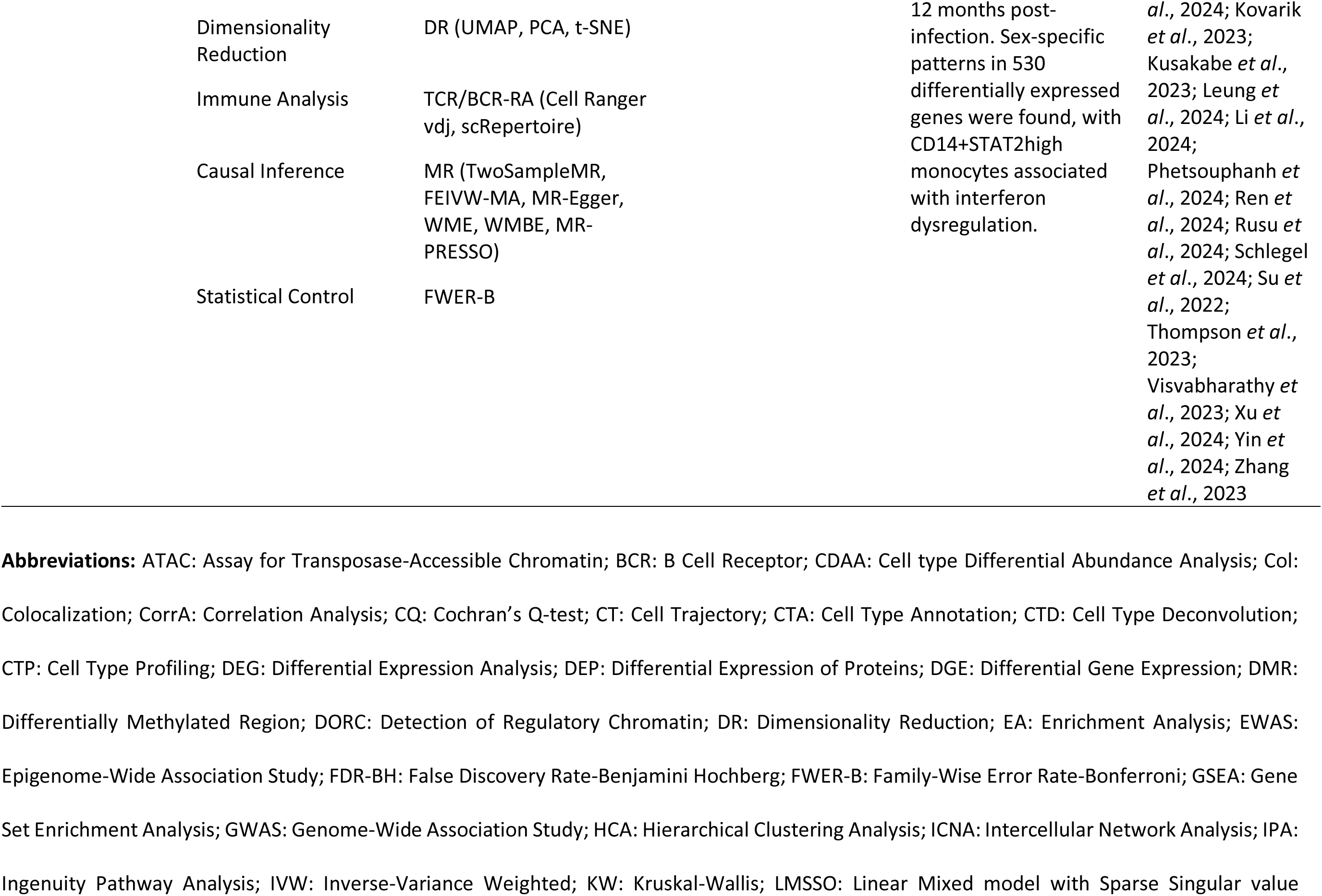
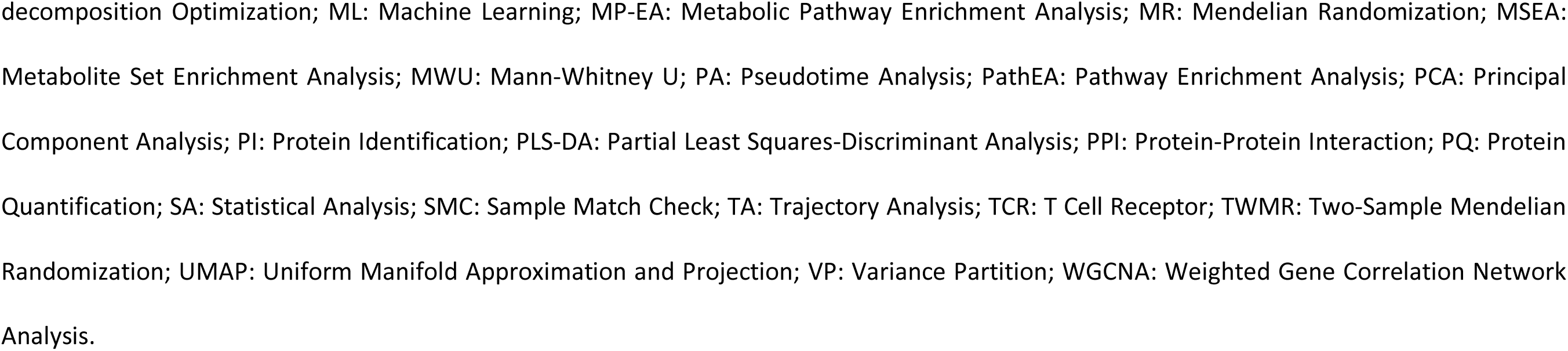
Computational Approaches and Biological Findings in Long COVID Research. This table provides an overview of computational approaches used in Long COVID research, including genome-wide and transcriptomic analyses, epigenomic profiling, proteomics, metabolomics, and multi-omics integration. We define computational methods as algorithmic approaches, statistical techniques, and software tools used to analyze, interpret, and derive insights from biological data—including data processing pipelines, statistical frameworks, machine learning algorithms, network analysis tools, and specialized bioinformatics software for omics data. The Analysis Categories column organizes these methods by analytical task, while Specific Methods & Tools lists the actual computational implementations. The table presents both these analytical methods and the resulting biological discoveries, highlighting key findings and relevant follow-up studies across multiple omics domains.

#### 3.2.1. Genomics

Genome-wide association studies (GWAS) have identified genetic variants associated with Long COVID. While multiple GWAS have focused on acute COVID-19 outcomes (COVID-19 Host Genetics Initiative, 2025), only two large-scale studies have systematically investigated genetic susceptibility to persistent symptoms as of March 2025 (Lammi *et al*., 2023; Deecke *et al*., 2025). These studies employed advanced statistical and computational methods to analyze genetic variations linked to Long COVID, revealing associations with immune-related pathways and complex disease traits.

Lammi *et al*. (2023) conducted the first GWAS meta-analysis for Long COVID across 24 studies from 16 countries, using Scalable and Accurate Implementation of GEneralized mixed model (SAIGE), a method designed to correct for population structure, sample relatedness, and case-control imbalance. Their analysis identified significant genetic variants in the FOXP4 locus, a gene previously implicated in lung function and immune regulation. Similarly, Chaudhary *et al*. (2024) performed a GWAS to identify genome-wide significant loci, including HLA-DQA1, ABO, and BPTF-KPAN2-C17orf58, which are involved in immune response and chronic inflammatory pathways.

To ensure robust findings, these studies applied meta-analytical frameworks to combine effect sizes and assess genetic associations while controlling for heterogeneity. Lammi *et al*. (2023) employed Fixed-effects Inverse-variance Weighted meta-analysis (IVW), Linear Mixed Model with Sparse Singular Value Decomposition Optimization (LMSSO), and Cochran’s Q-test (CQ) to evaluate the consistency of genetic effects across different populations. Beyond association studies, both GWAS leveraged Mendelian Randomization (MR) using LIMIX and Linkage Disequilibrium Score Regression (LDSC) to explore causal relationships between Long COVID and comorbid conditions such as chronic fatigue, fibromyalgia, and depression. Functional enrichment analysis (FEA) using FUMA, MAGMA, and STRING revealed shared genetic mechanisms between Long COVID and other post-viral syndromes (Lammi *et al*., 2023; Chaudhary *et al*., 2024).

To link genetic variants to biological function, secondary analyses integrated Expression Quantitative Trait Loci analysis (eQTL) with Genotype-Tissue Expression (GTEx) data, which confirmed increased FOXP4 expression in lung tissues. Colocalization analysis (COLOC) identified overlapping genetic signals between Long COVID and lung cancer risk. Single-cell RNA sequencing (scRNA-seq) further localized FOXP4 expression to alveolar and immune cell populations. Additional validation efforts included a Phenome-wide Association Study (PheWAS), which assessed the impact of identified variants across multiple clinical phenotypes. Genetic correlation analysis (GCA) using Linkage Disequilibrium Score Regression (LDSC) demonstrated a significant overlap between Long COVID susceptibility and inflammatory disease pathways (Lammi *et al*., 2023; Chaudhary *et al*., 2024).

A follow-up study (Pinero *et al*., 2025) expanded upon these findings by applying Two-Sample Mendelian Randomization (TSMR) and causal inference techniques such as Trans-ethnic Weighted Mendelian Randomization (TWMR) and Causal Transmission (CT) analysis. This study identified 32 causal genes (19 previously reported and 13 novel), which act as regulatory drivers influencing disease risk, progression, and stability. Enrichment analyses highlighted pathways linked to the SARS-CoV-2 response, viral carcinogenesis, cell cycle regulation, and immune function. Further analysis of other pathophysiological conditions revealed shared genetic factors across syndromic, metabolic, autoimmune, and connective tissue disorders. Moreover, this study proposed three distinct symptom-based subtypes of Long COVID based on genetic profiles, offering insights for more precise diagnosis and potential therapeutic interventions.

To explore regulatory elements influencing Long COVID risk loci, studies integrated DNase sequencing (DNase-seq) data from ENCODE, revealing that FOXP4 variants are located within active chromatin regions in the lungs. Statistical corrections, including Family-Wise Error Rate (FWER) control with Bonferroni correction, were applied to minimize false-positive associations (Lammi *et al*., 2023; Chaudhary *et al*., 2024).

#### 3.2.2. Epigenomics

Computational methods in epigenomics, which investigates heritable changes in gene functions without altering the DNA sequence, have been instrumental in uncovering the regulation of Long COVID. Several studies have analyzed DNA methylation patterns and chromatin accessibility to understand persistent immune and inflammatory alterations in affected individuals. Balnis *et al*. (2022) conducted a large-scale Epigenome-Wide Association Study (EWAS) to identify Differentially Methylated Regions (DMRs) in Long COVID patients. Their analysis employed statistical frameworks such as Linear Regression (LR) with empirical Bayes moderation and Enrichment Analysis (EA) using Gene Set Enrichment Analysis (GSEA) to determine biological pathways enriched in differentially methylated genes. Their findings revealed 71 persistent DMRs, 90% of which were in gene promoter regions, including immune and inflammatory regulators. Similarly, Nikesjo *et al*. (2025) identified persistent epigenetic alterations in Long COVID patients by analyzing DNA methylation profiles in neutrophil- and macrophage-enriched lung immune cell fractions, showing strong associations with immune response and cardiac function.

Lee *et al*. (2022) applied an alternative computational pipeline incorporating Differential Methylation Analysis (DMRcate) to evaluate methylation differences at single CpG sites. Their study identified significant hypomethylation in *IFI44L*, a gene linked to the innate immune response, along with hypermethylation at loci near PPT2 and RHOJ, which are associated with immune signaling. The researchers also applied Cell Type Composition (CTC) deconvolution methods to correct for cellular heterogeneity in blood samples, ensuring that observed methylation changes were not artifacts of shifts in immune cell proportions. Expanding on this, Nikesjo *et al*. (2025) employed DNA methylation profiling, with quality control and normalization performed via the SWAN method and batch effect assessment using ComBat. Their study incorporated PCA (Principal Component Analysis) for primary analyses, identifying differentially methylated CpG sites through linear models that assessed longitudinal DNAm changes. Secondary computational analyses using STRING, KEGG, and Gene Ontology (GO) confirmed functional enrichment of differentially methylated genes, with pathway analyses highlighting persistent immune dysregulation, Wnt signaling, and circadian rhythm involvement.

Additional large-scale studies have expanded the understanding of Long COVID epigenetics. A study utilizing Whole Genome Methylation Sequencing (WGMS) identified 39 DMRs distinguishing Long COVID patients from controls. These DMRs were linked to quality-of-life impairment, with motif analysis showing enrichment of circadian rhythm regulators (Balnis *et al*., 2022). Similarly, Nikesjo *et al*. (2025) identified persistent hypomethylation and hypermethylation at key immune loci in lung immune cells, with DNA methylation changes correlating with pathways involved in cardiac function, Wnt signaling, and circadian rhythm. Their validation approach included independent datasets and correlation with a symptom-physiology score, reinforcing the robustness of their findings.

Moreover, epigenetic drift and accelerated aging were observed in patients and assessed via Horvath’s epigenetic clock. Functional enrichment analyses further highlighted dysregulation in immune-related pathways (Balnis *et al*., 2024). A third study integrated global DNA methylation analysis, telomere length measurement, and mitochondrial DNA copy number assessment to identify six Long COVID symptom clusters. PCA and K-means clustering separated patient groups, while regression models explored biological predictors. Functional enrichment analyses (STRING, ROC analysis) identified associations between telomere shortening, mitochondrial dysfunction, and symptom severity, particularly post-exertional malaise-fatigue. Expanding on this, Nikesjo *et al*. (2025) demonstrated that DNA methylation changes in lung immune cells were significantly associated with symptoms and physiological measures, providing further support for a link between immune epigenetics and disease severity.

To explore the functional implications of these epigenetic modifications, all studies integrated chromatin accessibility data from DNase sequencing (ENCODE) to map regulatory regions affected by methylation alterations. Additionally, MR techniques were employed to infer causal relationships between identified epigenetic modifications and Long COVID susceptibility (Balnis *et al*., 2022; Balnis *et al*., 2024; Lee *et al*., 2022; Polli *et al*., 2025). Nikesjo *et al*. (2025) further extended this by analyzing DNA methylation changes in neutrophil- and macrophage-enriched lung immune cells, demonstrating persistent DNAm patterns linked to immune regulation and cardiac function, which could serve as potential targets for epigenetic-modulating drugs.

Furthermore, statistical controls such as the Benjamini-Hochberg False Discovery Rate (FDR-BH) and Family-Wise Error Rate (FWER-B) correction were applied to mitigate false-positive findings. These computational approaches collectively suggest that persistent DNA methylation changes in key immune genes may contribute to Long COVID pathophysiology, highlighting potential targets for therapeutic intervention (Balnis *et al*., 2022; Balnis *et al*., 2024; Lee *et al*., 2022; Polli *et al*., 2025; Nikesjo *et al*., 2025).

#### 3.2.3. Transcriptomics

Transcriptomic analyses provide insights into gene expression changes associated with Long COVID, identifying molecular pathways linked to immune dysfunction, inflammation, and tissue remodeling. These studies employ bulk RNA sequencing (RNA-seq) and single-cell RNA sequencing (scRNA-seq) to characterize transcriptional differences between affected individuals and controls, each requiring distinct computational methodologies.

Bulk RNA sequencing studies primarily rely on Differential Gene Expression (DGE) analysis to identify genes with significant expression changes. Multiple studies have implemented DGE to examine transcriptional alterations in Long COVID, revealing dysregulated immune pathways and inflammatory markers (Blankestijn *et al*., 2024; García-Hidalgo *et al*., 2022; Greene *et al*., 2024; Mehta *et al*., 2024; Ryan *et al*., 2022). RNA-seq data processing involved read alignment and transcript quantification, with methods such as HiSAT2 for spliced read alignment and edgeR for statistical modeling of gene expression. Variance partition analysis (VPA) was applied to assess the contribution of technical and biological factors to transcriptional variability, ensuring robust identification of differentially expressed genes.

Building upon Greene *et al*.’s (2024) brain-related research, follow-up studies employed increasingly sophisticated computational approaches. One study analyzed RNA-seq data to identify blood-brain barrier (BBB) dysfunction markers, revealing BBB disruption, persistent inflammation, and mitochondrial pathway dysregulation correlating with cognitive impairment (Dhariwal *et al*., 2024). Another follow-up analyzed 22 PBMC samples using GEO2R with limma and GEOquery packages, protein-protein interaction networks via STRING and Cytoscape, and functional pathway enrichment with Metascape. This approach identified 250 differentially expressed genes in brain fog patients, including *SMAD3* (neuroinflammation), *PF4* (blood clotting), *CXCL5* (BBB damage), and *TNFAIP3* (cytokine regulation), confirming systemic inflammation and BBB dysfunction mechanisms (Chakraborty *et al*., 2024a). A third follow-up focused on *FOXP4*, analyzing four RNAseq datasets with population genetic analysis and evolutionary analysis, resulting in identifying potential *FOXP4* inhibitors through molecular docking (Gupta *et al*., 2024).

Ryan *et al*.’s (2022) work on long-term immune changes generated multiple follow-up studies. One analyzed GEO datasets using WGCNA algorithm and immune infiltration analysis (CIBERSORT/ssGSEA/xCell) to characterize transcriptional patterns in Long COVID (Hou *et al*., 2025b). A transcriptomic meta-analysis of three RNA-seq datasets comprising 135 samples revealed 530 differentially expressed genes and sex-specific expression patterns with ribosomal proteins showing opposite regulation between males and females (Rusu *et al*., 2024). Another study compared Long COVID with Chronic Thromboembolic Pulmonary Hypertension, identifying three shared key hub genes (*DNAJA1*, *NDUFA5*, *SLC2A14*) and pathways related to immune modulation, oxidative stress, and metabolism (Li *et al*., 2025a). A meta-analysis comparing Long COVID with autoimmune diseases found that while interferon signatures were upregulated in severe COVID-19 and autoimmune diseases, they were downregulated in Long COVID, suggesting interferon exhaustion as a potential driver of pathogenesis (Gómez-Carballa *et al*., 2024).

Additional follow-ups on Ryan’s work (Ryan *et al*., 2022) included microbiome re-annotation of RNA-Seq data revealed blood microbiome disturbances at 12 weeks post-infection, with increased Bacillota/Bacteroidota ratio potentially linked to olfactory dysfunction (Wang *et al*., 2024). One study compared Long COVID with ME/CFS, identifying nine common genes (including *CXCL8*, *B2M*, *SOD1*, *BCL2*) and constructed a transcription factor-miRNA coregulatory network (Lv *et al*., 2022). A Polygenic Risk Score (PRS) analysis demonstrated a genetic link between Long COVID risk and early-differentiated CD4+ T cell distributions in healthy individuals (Deecke *et al*., 2024).

Pathway enrichment and functional annotation analyses characterized transcriptomic changes in Long COVID using Gene Ontology (GO) enrichment, KEGG pathway analysis, and dimensionality reduction techniques (t-SNE, UMAP). These approaches identified immune system activation, dysregulated interferon signaling, and coagulation pathways. Hierarchical clustering identified patient subgroups, and STRING analysis facilitated pathway enrichment interpretation (Ryan *et al*., 2022).

Single-cell RNA sequencing provided higher-resolution analysis of Long COVID through cell type annotation algorithms (Seurat, Leiden clustering), trajectory inference methods, and cell-cell communication analysis. Gillrie *et al*.’s (2023) immune profiling work was extended through scRNAseq analysis of MIS-A patient samples, revealing novel neutrophil populations expressing MMP9/CD177/IGHA1/XBP1, unique plasmablast B-cell subsets, and coagulation-dependent microvascular dysfunction, establishing immunothrombosis as a key pathogenic mechanism.

Follow-ups on Yao *et al*.’s (2023) TGF-beta research conducted scRNA-seq on bronchoalveolar lavage cells and PBMCs from COVID-19 convalescents, identifying altered immune landscapes in respiratory Long COVID with increased monocyte-derived alveolar macrophages correlating with impaired lung function. They found IFN-γ as a key driver produced by tissue-resident T cells, suggesting JAK inhibitor Baricitinib as a potential therapeutic based on IFN-γ pathway involvement (Li *et al*., 2023).

Finlay *et al*.’s (2022) smell loss research used scRNA-seq analysis of olfactory epithelial biopsies, employing Scanpy and scvi-tools for preprocessing and Wilcoxon rank sum tests for differential expression. This revealed persistent T cell-mediated inflammation long after viral clearance, with enriched interferon-γ expressing CD8+ T cells, altered myeloid populations, and reduced olfactory sensory neurons, establishing a mechanistic basis for persistent smell loss.

Vaivode *et al*. (2024) conducted scRNA-seq on peripheral blood cells from Long COVID patients before and after herbal therapy, using BD Rhapsody WTA pipeline and Seurat for analysis. They identified persistent T cell activation, altered monocyte subtypes, changes in neutrophil activity linked to prolonged inflammation, and transcriptional heterogeneity in treatment responses.

These computational approaches collectively revealed key findings about Long COVID pathophysiology: alterations in immune-related genes emphasize persistent immune activation; neurological studies confirmed BBB disruption; lung-specific analyses identified fibrosis-associated changes; immune profiling demonstrated prolonged inflammation and altered T-cell responses; and spatial transcriptomics mapped how IFN and TNF-mediated macrophage activation leads to IL-1β release and fibrosis. Additional insights came from plasma microRNA profiling, identifying miR-9-5p and miR-486-5p as regulators of angiogenesis and inflammatory pathways associated with pulmonary dysfunction, while airway mucosa transcriptomic profiling revealed neutrophil infiltration and epithelial dysfunction with upregulated neutrophil activation pathways, mucin gene expression, and immune dysregulation in Long COVID patients.

#### 3.2.4. Proteomics

Proteomic analyses, which study the complete set of proteins expressed in biological samples, have applied different computational methods for Long COVID from statistical assessments of Differentially Expressed Proteins (DEPs) to ML-driven biomarker discovery.

Primary analyses focused on identifying DEPs using statistical methods, including t-tests, Mann-Whitney U tests, and linear regression. To ensure biological relevance, fold-change thresholds were applied. Multiple studies (Li *et al*., 2022; Peppercorn *et al*., 2023; Pretorius *et al*., 2021) implemented these approaches, revealing immune dysregulation and inflammatory protein signatures associated with Long COVID. Data processing incorporated specialized mass spectrometry techniques, including Data-Dependent Acquisition (DDA) and Data-Independent Acquisition (DIA), which captured comprehensive protein profiles from complex biological samples. Olink Proteomics panels were also used for targeted quantification (Liew *et al*., 2024).

Secondary computational analyses employed various approaches. Enrichment Analysis (EA) and Protein-Protein Interaction (PPI) network analysis identified key functional pathways involved in inflammation and metabolic dysregulation. Network analysis tools such as STRING, GSEA, and Weighted Gene Co-expression Network Analysis (WGCNA) provided further insights into protein relationships and immune-mediated processes (Grote *et al*., 2024; Iosef *et al*., 2023). Hierarchical and k-means clustering techniques were applied to group proteins and patient samples by similarity (Zoodsma *et al*., 2022).

Advanced proteomic profiling enabled pathway-level insights into disease pathophysiology. Studies using SureQuant PQ500 panels identified differential abundance of plasma proteins linked to platelet activation, extracellular matrix remodeling, and immune dysfunction. Functional enrichment analyses mapped immune activation pathways, coagulation abnormalities, and lipid metabolism shifts, highlighting their persistence in Long COVID patients (Suski *et al*., 2024). Inflammatory marker profiling using Olink multiplex panels further demonstrated TNF-a signaling and complement system dysregulation in post-COVID-19 cases (Patel *et al*., 2023).

Single-cell proteomics and spatial transcriptomics approaches refined these findings by elucidating tissue-specific protein changes. Analyses of bronchoalveolar lavage fluid (BAL) using DIA-MS and PEA proteomic panels identified ongoing lung repair processes and immune activation up to 9 months post-infection (Kanth *et al*., 2024). Similarly, plasma-derived neuronal extracellular vesicle (nEV) profiling highlighted elevated neurodegenerative markers (Aß42, pTau181, TDP-43), suggesting persistent neuronal injury and cognitive impairment in Long COVID patients (Tang *et al*., 2024).

A computational study explored potential therapeutic targets. Immunomodulatory pathways were highlighted as key drivers of persistent inflammation, with complement activation and myeloid-driven immune responses central to Long COVID symptomatology (Mohammed *et al*., 2024).

Finally, proteomic studies have contributed to understanding exercise-induced molecular changes in Long COVID patients. A recent DIA-MS study analyzed plasma proteins before and after an 8-week exercise program, revealing reductions in inflammatory markers (S100A8, CST3) and oxidative stress-related proteins (Chowdhury *et al*., 2024).

#### 3.2.5. Metabolomics

Computational approaches in metabolomics have revealed critical biochemical alterations in Long COVID, providing insights into disease mechanisms through systematic analysis of metabolites. These methods ranged from fundamental statistical analyses to advanced classification techniques.

Primary analysis employed several statistical approaches to identify significantly altered metabolites in Long COVID patients. Univariate statistical tests, such as the Mann-Whitney U test and PLS-DA regression, were applied to determine differential metabolite abundance, while correlation analyses (Pearson’s and Spearman’s) were used to link metabolite levels with clinical symptoms (Holmes *et al*., 2021; López-Hernández *et al*., 2023; Mikuteit *et al*., 2023). These approaches identified 19 Differentially Expressed Metabolites (DEMs) in saliva, including reductions in creatine, Gly Pro Lys, and calenduloside G methyl ester, suggesting metabolic dysfunction (Machado *et al*., 2024).

Pattern recognition methods enabled metabolomic profile classification. PCA and hierarchical clustering were used to detect metabolic subgroup differences, while pathway enrichment analysis (STRING, HMDB, Metlin Library) identified metabolic pathways affected in Long COVID patients. Studies found disruptions in mitochondrial function, amino acid metabolism, and lipid dysregulation, with 46 metabolites differentiating Long COVID from recovered patients (Martinez *et al*., 2024). Altered mitochondrial metabolism was indicated by increased TCA cycle activity and decreased ceramide levels, linking these metabolic shifts to persistent immune activation. Other metabolomic studies identified disruptions in fatty acid oxidation, with elevated pyruvate, 3-hydroxyisovaleric acid, and cystine levels in serum samples, further supporting mitochondrial dysfunction (Anda *et al*., 2024).

Advanced metabolomic analyses incorporated clustering algorithms, such as HDBSCAN and K-means, which revealed metabolomic subgroups in Long COVID cohorts, supporting the heterogeneity of the condition. Further computational integration with STRING and MetaboAnalyst allowed functional annotation of differentially abundant metabolites, correlating them with immune dysfunction and mitochondrial dysregulation (Mendes *et al*., 2024). Studies employing Proton Nuclear Magnetic Resonance (1H NMR) spectroscopy and Liquid Chromatography-Mass Spectrometry (LC-MS) further identified metabolic shifts over time, revealing alterations in glucose metabolism, amino acid imbalances, and mitochondrial dysfunction in Long COVID patients (Ansone *et al*., 2024). Additionally, metabolomic studies integrated gut microbiota data, uncovering metabolic dysbiosis in post-COVID individuals, with increased succinic and fumaric acids linking inflammation and mitochondrial impairment (Sorokina *et al*., 2023).

Key metabolomic studies found consistent metabolic disruptions across multiple datasets and experimental approaches. Dysregulation of amino acid metabolism, mitochondrial dysfunction, and lipid metabolism were persistent features in Long COVID patients. Validation approaches included Bonferroni correction, cytokine profiling, and independent cohort comparisons to confirm metabolic changes. Metabolomic disturbances were also correlated with inflammatory markers and immune responses, further strengthening findings related to Long COVID pathophysiology (Anda *et al*., 2024; Ansone *et al*., 2024; Holmes *et al*., 2021; López-Hernández *et al*., 2023; Machado *et al*., 2024; Martinez *et al*., 2024; Mikuteit *et al*., 2023; Sorokina *et al*., 2023)

#### 3.2.6. Multi-omics

Integrating multiple molecular data types through computational methods has provided comprehensive insights into Long COVID pathology. Multi-omics approaches combine different molecular measurements to understand disease mechanisms at multiple biological levels.

Primary analyses in multi-omics studies centered on differential expression analysis across multiple data layers. Variance partition analysis was employed to account for technical variables, while statistical frameworks such as CIBERSORTx were used for cell type-specific differential expression analysis, allowing identification of transcriptomic differences in Long COVID patients (Thompson *et al*., 2023; Eaton-Fitch *et al*., 2024). A follow-up study based on Thompson *et al*., 2023 (Ren *et al*., 2023) analyzed whole blood RNA-seq data to identify genes associated with Long COVID outcomes, revealing key genes including *CCDC18* (involved in viral RNA regulation), *ADARB2* (related to inflammation through RNA editing), and immune-related genes *CXCL8*, *IL4R*, and *IGHM*. Their analysis highlighted immune cell infiltration and inflammatory signaling as key mechanisms in Long COVID. Another follow-up study on the same dataset revealed that frail COVID-19 patients showed higher rates of Long COVID symptoms, with 477 differentially expressed genes enriched in inflammatory pathways including AGE-RAGE signaling, and identified *NCAPG*, *MCM10*, and *CDC25C* as hub genes involved in cell division regulation with potential as diagnostic biomarkers (Li *et al*., 2023).

Further analysis of Thompson *et al*., 2023 examined mitochondrial dysfunction in Long COVID by focusing on nuclear-encoded mitochondrial (NEM) genes. This approach identified 380 unique NEM differentially expressed genes in Long COVID patients compared to only 14 in convalescent individuals, with enriched pathways related to mitochondrial membrane organization, complex assembly, and protein transport (Silverstein *et al*., 2024).

A study applied interaction models to uncover gene expression signatures specific to plasma cells, with distinct pulmonary and miscellaneous clusters dependent on anti-spike antibody titers. Further analyses using ELISA quantified anti-spike antibodies, integrating immune response data with transcriptomic profiles (Eaton-Fitch *et al*., 2024).

Similar findings emerged from analysis of Su *et al*., 2022, which examined single-cell transcriptomics from three independent cohorts. This study discovered that SARS-CoV-2 infection leads to prolonged alteration of gene expression in circulating immune cells lasting at least 2 months after symptom onset, particularly affecting NEM genes. Gene network analysis suggested a central role for NF-κB transcription factor in regulating observed transcriptional changes, with severe/critical COVID-19 patients reporting Long COVID symptoms showing lower time-adjusted IL2-AIS scores, suggesting more pronounced inflammatory responses (Zhang *et al*., 2023).

Different computational techniques for multi-omics integration were used to combine molecular data from different platforms. Single-cell multi-omics approaches integrated scRNA-seq with chromatin accessibility data (snATAC-seq) and protein measurements (CITE-seq) to reveal coordinated cellular responses in Long COVID (Cheong *et al*., 2023). A follow-up study on Cheong’s dataset revealed lasting transcriptional changes in hematopoietic stem and progenitor cells from severe COVID-19 patients during recovery, particularly in granulocyte myeloid progenitors. These cells showed increased expression of genes encoding C-type lectin receptors and downstream signaling molecules that persisted up to 12 months post-infection, suggesting a mechanism whereby gut fungal dysbiosis during acute infection leads to lasting immune alterations in bone marrow progenitors (Kusakabe *et al*., 2023).

Multiple follow-up studies of Yin *et al*.’s (2024) dataset provided additional insights. One analysis identified 16 key genes with distinct functional roles including *IGHG4*, *NAIPP4*, and *TRAV8-5* involved in adaptive immunity; *MDGA1* and *KNDC1* associated with neurological symptoms; and several genes linked to transcriptional regulation and immune dysregulation (Hou *et al*., 2025b). Another study analyzed scRNA-seq data from peripheral blood samples of non-elderly female participants, identifying a distinctive CD14+STAT2 high monocyte subset in Long COVID patients showing increased expression of *STAT2* and *IRF9* transcription factors, suggesting persistent type I interferonopathy as a key mechanism (Xu *et al*., 2024).

A meta-analysis identified 530 differentially expressed genes in Post-COVID Condition, with pathways implicated in cell cycle processes, immune dysregulation, and histone modifications. Sex-specific analyses revealed distinct expression patterns between males and females, with male PCC primarily associated with neuro-cardiovascular disorders, while females exhibited more diverse conditions with higher incidence of respiratory conditions (Rusu *et al*., 2024). Additionally, pathway enrichment tools such as GO-Figure!, STRING, and WGCNA were used to analyze shared gene expression profiles and metabolic shifts. Studies identified inflammatory and platelet degranulation, and lipid metabolism alterations by linking RNA-seq and LC-MS/MS datasets (Wei *et al*., 2024).

An important study on interferon (IFN) gene expression patterns analyzed five cohorts of acute COVID-19 and Long COVID patients, comparing them to expression patterns in three autoimmune diseases. While IFN signatures showed strong upregulation in severe COVID-19 patients and autoimmune diseases, the signals decayed over time from symptom onset and were diminished in Long COVID patients. IFN-related pathway analysis indicated overactivation in autoimmune diseases and severe COVID-19, while these pathways were mostly inactivated or downregulated in Long COVID, suggesting cytokine exhaustion as a possible driver rather than sustained activation (Chakraborty *et al*., 2024a).

Advanced statistical approaches were applied to stratify Long COVID subgroups based on multi-omics signatures. PCA, PLS-DA, and Welch’s t-test identified proteomic and metabolomic shifts distinguishing Long COVID from recovered individuals. Hierarchical clustering techniques uncovered novel patient endotypes linked to persistent immune activation and coagulation abnormalities (Wang *et al*., 2023). Additionally, studies integrating RNA-seq and proteomic data identified a Long COVID subgroup with heightened neutrophil degranulation and inflammatory responses (Lin *et al*., 2024).

These multi-omics studies revealed persistent immune dysregulation, metabolic shifts, and platelet activation as defining features of Long COVID. Validation strategies included independent dataset replication, correlation with clinical outcomes, and comparison with external single-cell and proteomic datasets. Longitudinal tracking of multi-omics changes has further highlighted sustained alterations in inflammatory signaling, lipid metabolism, and immune responses (Lin *et al*., 2024; Wang *et al*., 2023), with particular emphasis on mitochondrial dysfunction, altered T cell responses, and sex-specific molecular signatures as key factors in Long COVID pathophysiology.

### 3.3. Long COVID Classification and Prediction

Understanding and predicting Long COVID subtypes and evaluating the risk of developing this condition are crucial for optimizing treatment strategies and improving patient outcomes. This section introduces the methodologies and findings related to the classification and prediction of Long COVID manifestations (Table 2).

**Table 2.**
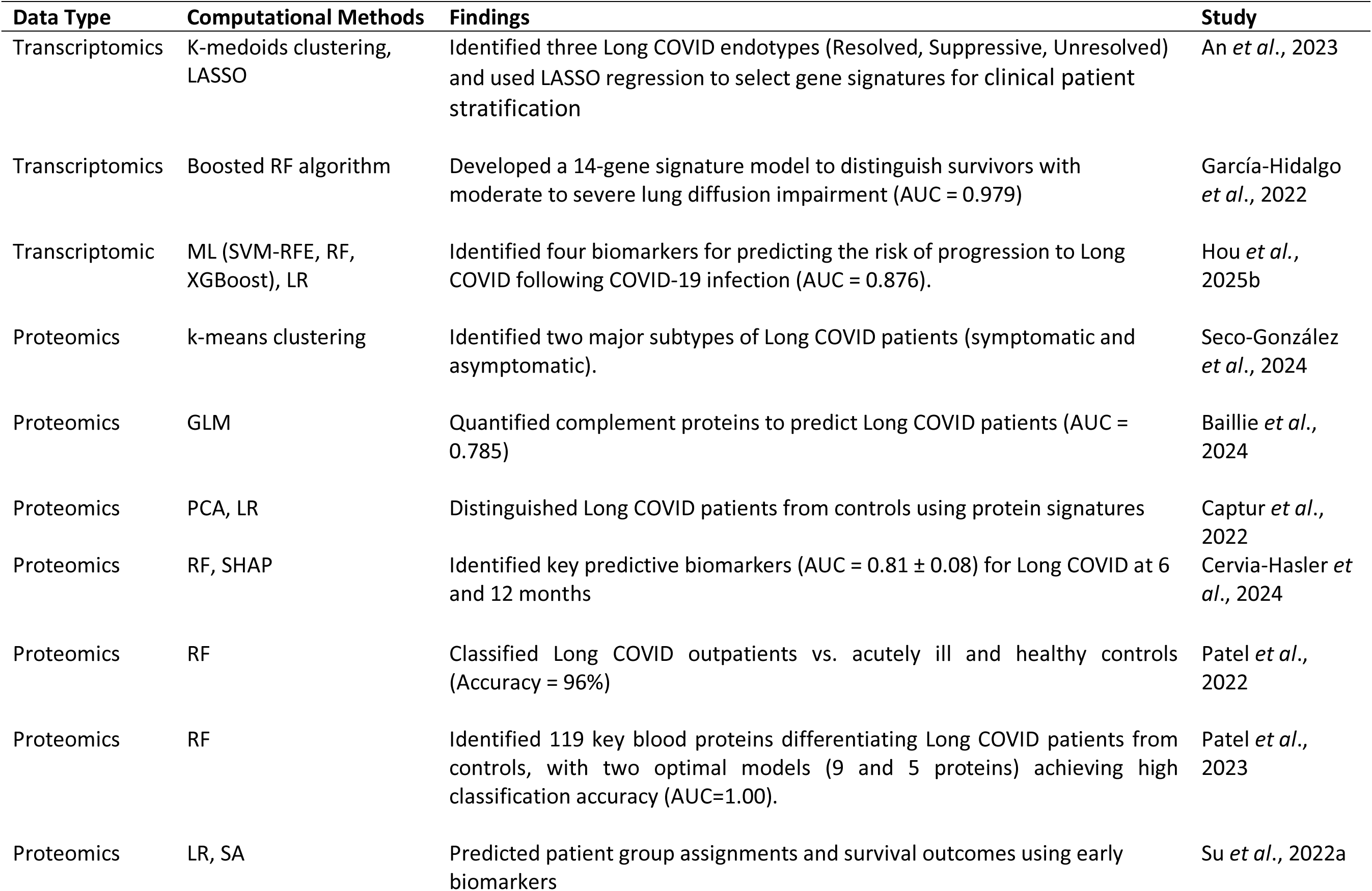
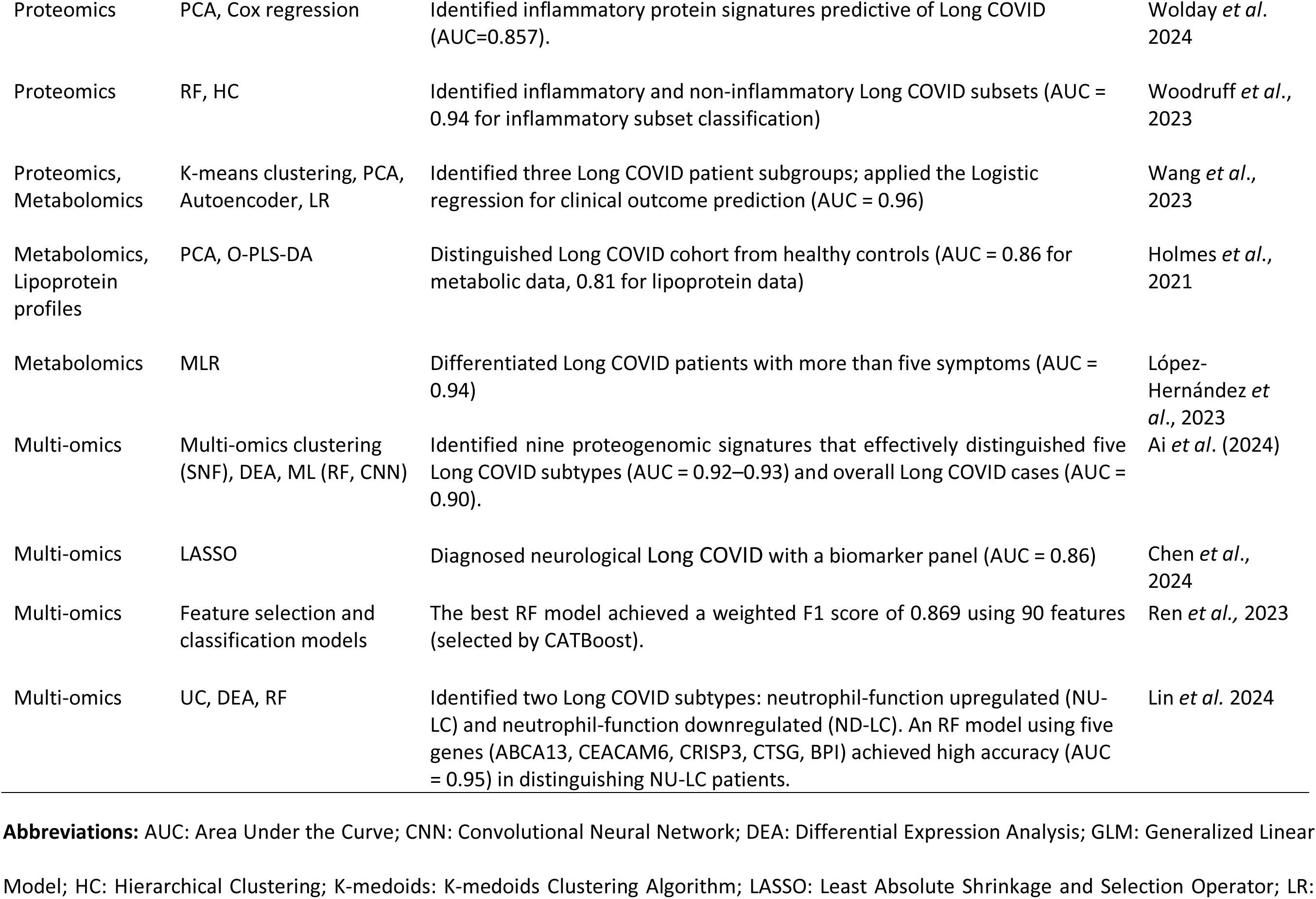
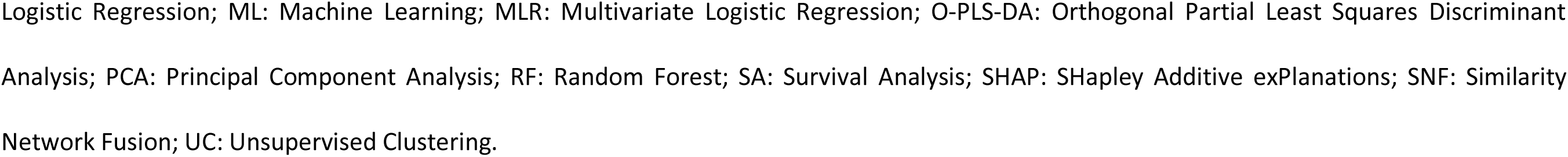
Computational Methods for Long COVID Classification and Prediction.

Unsupervised clustering techniques have been widely applied to classify Long COVID subtypes based on molecular signatures. An *et al*. (2023) used K-medoids clustering to identify three endotypes, resolved, suppressive, and unresolved, each associated with distinct symptom trajectories and gene expression profiles. Similarly, Wang *et al*. (2023) employed k-means clustering on autoencoder-derived features to define three clinically distinct Long COVID groups, highlighting persistent metabolic and immune dysregulation.

Proteomic data has also facilitated subtype identification. Seco-González *et al*. (2024) applied k-means clustering to distinguish symptomatic from asymptomatic Long COVID patients and developed a proteomic severity scale. Woodruff *et al*. (2023) used hierarchical clustering to delineate inflammatory (infLC) and non-inflammatory (niLC) subtypes, revealing immunologic heterogeneity.

Beyond clustering, dimensionality reduction methods have enhanced classification. Captur *et al*. (2022) applied Principal Component Analysis (PCA) to proteomic data, successfully distinguishing Long COVID patients from controls. Holmes *et al*. (2021) combined Principal Component Analysis (PCA) with Orthogonal Partial Least Squares-Discriminant Analysis (O-PLS-DA), achieving Area Under the Curve (AUC) values of 0.86 and 0.81 for metabolic and lipoprotein profiling, respectively. Ai *et al*. (2024) integrated multi-omics clustering via Similarity Network Fusion (SNF), stratifying Long COVID into five molecular subtypes. Wang *et al*. (2023) combined Principal Component Analysis (PCA) with autoencoder-based feature extraction for cluster refinement, while Cervia-Hasler *et al*. (2024) incorporated SHapley Additive exPlanations (SHAP) explainability techniques for biomarker selection.

Supervised learning has been instrumental in predicting Long COVID subtypes, risk, and outcomes. García-Hidalgo *et al*. (2022) trained a boosted Random Forest (RF) classifier to predict lung function impairment (Diffusing capacity of the Lungs for Carbon Monoxide (D_LCO) <60%) with an Area Under the Curve (AUC) of 0.979 using a reduced gene signature. Hou *et al*. (2025b) applied Support Vector Machine-Recursive Feature Elimination (SVM-RFE), Random Forest (RF), and eXtreme Gradient Boosting (XGBoost) for feature selection, identifying key biomarkers (*CEP55*, *CDCA2*, *MELK*, *DEPDC1B*) with an Area Under the Curve (AUC) of 0.876 for Long COVID risk prediction.

Several studies have leveraged proteomic and metabolomic data for predictive modeling. Patel *et al*. (2024) applied Random Forest (RF) modeling with a feature selection method to plasma proteomics, developing models that correctly classified Long COVID patients (Area Under the Curve (AUC) = 1.00). Cervia-Hasler *et al*. (2024) used Random Forest (RF) classification with SHapley Additive exPlanations (SHAP) analysis to identify predictive biomarkers, achieving an Area Under the Curve (AUC) of 0.81. Chen *et al*. (2024) integrated Least Absolute Shrinkage and Selection Operator (LASSO) regression to develop a biomarker panel (ST1A1, sphinganine, 7,8-dihydro neopterin) for diagnosing neuro-Long COVID (Area Under the Curve (AUC) = 0.86). Baillie *et al*. (2024) employed a generalized linear model with complement protein markers (iC3b, Terminal Complement Complex (TCC), Ba, C5a) to predict Long COVID severity (Area Under the Curve (AUC) = 0.785).

Predictive models have also been used for survival and symptom progression. Su *et al*. (2022b) trained logistic regression models on plasma protein data to predict Long COVID endotypes and survival outcomes. Wolday *et al*. (2024) combined clustering, differential expression analysis, and Cox proportional hazard models to identify inflammatory risk markers (Signaling Lymphocytic Activation Molecule Family member 1 (SLAMF1), Tumor Necrosis Factor (TNF), Interleukin 15 Receptor Alpha (IL15RA), Interleukin 18 (IL18), C-X-C Motif Chemokine Ligand 10 (CXCL10)) and protective factors (TNF-Related Activation-Induced Cytokine (TRANCE)). Ren *et al*. (2023) integrated multiple Machine Learning (ML) algorithms (Least Absolute Shrinkage and Selection Operator (LASSO), Light Gradient Boosting Machine (LightGBM), Random Forest (RF), eXtreme Gradient Boosting (XGBoost)) with Synthetic Minority Oversampling Technique (SMOTE) to develop a robust Long COVID risk prediction model, achieving a weighted F1 score of 0.869.

A range of unsupervised and supervised computational approaches have been applied to classify and predict Long COVID subtypes, risk, and outcomes. Clustering methods have been effective in identifying distinct phenotypes, while supervised learning has enabled accurate classification and prediction using gene, proteomic, and metabolomic markers. However, challenges such as small sample sizes, case-definition inconsistencies, and the need for external validation persist. Future research should focus on large-scale, multi-omic studies with independent validation to enhance model reliability and clinical applicability.

### 3.4. Long COVID Treatment

There is lack of treatment for Long COVID. The current efforts primarily focus on symptom management and quality-of-life improvement (Shaffer, 2022). Managing Long COVID symptoms needs a holistic, multidisciplinary approach involving structured primary care visits, rehabilitation services, and patient empowerment (Sisó-Almirall *et al*., 2021).

Evidence supports SARS-CoV-2 vaccination to help prevent Long COVID, and it is recommended that Long COVID patients adhere to standard vaccination schedules (Ceban *et al*., 2023). Early antiviral therapy has also been associated with a reduced risk of Long COVID and lower hospitalization or death rates, suggesting that early antiviral treatment is advisable for at-risk individuals (Choi *et al*., 2023). However, evidence regarding the impact of antiviral treatments during acute COVID-19 on subsequent development of Long COVID remains conflicting (Y. Su *et al*., 2022b). Some studies suggest that Remdesivir or Nirmatrelvir/Ritonavir could exert a protective effect for Long COVID, while a limited number have indicated possible benefits of medications such as Dexamethasone or Metformin, though more research is needed (Fernández-de-Las-Peñas *et al*., 2023). Overall, the scarcity of evidence-based treatment guidelines and approved therapies continues to pose significant challenges for healthcare providers (Schrimpf *et al*., 2022).

Computational methods play a crucial role in identifying therapeutic strategies for Long COVID by leveraging biomarker discovery, pathway analysis, gene expression modulation, and patient subtyping. These approaches facilitate drug repurposing and precision medicine, targeting the diverse pathophysiological mechanisms underlying the condition.

Drug-target interaction analyses have identified potential therapeutics for Long COVID. For instance, TUBB4A, linked to severe lung diffusion impairment, was found to be targeted by six FDA-approved drugs (García-Hidalgo *et al*., 2022). Similarly, targeting chemokine receptors (e.g., CXCR4) and IL6 has been suggested to reduce immune infiltration and persistent inflammation, with ongoing trials investigating antiviral therapies like nirmatrelvir-ritonavir (Yin *et al*., 2024). Additionally, anticoagulants such as heparin, hirudin, and activated protein C (APC) have been proposed for vascular dysfunction prevention due to their endothelial-protective effects (Gillrie *et al*., 2023).

Pathway-based analyses have highlighted several therapeutic targets. HIF-1 pathway inhibitors and AHR antagonists have shown potential in modulating immune and metabolic dysregulation in Long COVID (Iosef *et al*., 2023; Guo *et al*., 2023). Using Ingenuity Pathway Analysis (IPA), key inflammatory pathways and upstream regulators were identified, with glucocorticoids such as budesonide and fluticasone propionate emerging as potential treatments to mitigate airway inflammation (Gerayeli *et al*., 2024). Furthermore, targeting the terminal complement pathway has been proposed to address immune dysregulation (Cervia-Hasler *et al*., 2024). Comprehensive cellular and molecular analyses suggest that inhibiting specific immune pathways and localized drug delivery could offer effective therapeutic strategies (Ryan *et al*., 2022; Finlay *et al*., 2022; Cheong *et al*., 2023).

Molecular profiling has identified potential interventions based on observed alterations in gene expression and metabolomic changes. Sarcosine and serine supplementation have been proposed to counteract metabolic disruptions observed in Long COVID patients (Saito *et al*., 2024). Additional candidates include taurine, citrulline, glutamine, antioxidants (e.g., N-acetylcysteine, NAD+), and arginine; however, these remain to be clinically validated (López-Hernández *et al*., 2023). Moreover, reverse gene expression screening has been used to identify compounds capable of modulating disease-associated transcriptional changes, providing further therapeutic avenues (Lv *et al*., 2022).

Integrated computational approaches combining drug-target interactions, pathway perturbation effects, and gene expression modulation have identified potential drug candidates. For example, enrichment analysis using DSigDB identified drugs capable of reversing disease-associated molecular signatures in Long COVID patients (Lv *et al*., 2022).

Subtyping patients with distinct Long COVID phenotypes may facilitate specific interventions. Yong & Liu (2022) conducted a narrative review that characterized six subtypes of Long COVID based on existing literature, each presenting unique symptoms and pathophysiological mechanisms. Clarifying these subtypes could inform targeted treatment design and advance medical and public health efforts to reduce Long COVID’s substantial health and economic burdens. Similarly, Woodruff *et al*. (Woodruff *et al*., 2023) highlights the diagnostic and therapeutic potential of defining a clearly identifiable disease subtype, with implications for ongoing investigations into immunomodulatory therapy as a Long COVID treatment.

Research into Long COVID continues to evolve, and new treatment options may emerge in the future. Identifying potential therapeutics using omics data constitutes a preliminary step toward clinical trials. Despite increasing research efforts, most tested drugs have not demonstrated high efficacy in RCTs. Given the complex nature of Long COVID, ongoing studies are essential to refine subtype-specific interventions and to advance the understanding of its pathobiology.

## 4. Future Challenges and Directions

Despite recent advances, each omics domain in Long COVID research faces unique challenges. We found that a common barrier across all omics studies is the limited cohort size and diversity, which restricts statistical power and generalizability (Balnis *et al*., 2022; García-Hidalgo *et al*., 2022; Holmes *et al*., 2021; Lammi *et al*., 2023; Peppercorn *et al*., 2023). Collaborative multi-center studies and consortia are needed to gather larger, more diverse patient cohorts (COVID-19 Host Genetics Initiative, 2025; Munblit *et al*., 2022).

In genomics, studies have begun to identify genetic risk factors for Long COVID (Gupta *et al*., 2024; Lammi *et al*., 2023), though many analyses remain underpowered and poorly represent non-European ancestries (Deecke *et al*., 2025). From our perspective, future research should include diverse populations and whole-genome sequencing, while developing polygenic risk scores and integrating Mendelian randomization approaches to find causal genetic contributors (Pinero *et al*., 2025).

Epigenomic studies in Long COVID currently cover only a fraction of the epigenome, focusing mostly on DNA methylation (Balnis *et al*., 2022; Lee *et al*., 2022). Our analysis indicates that more comprehensive approaches including histone modifications, chromatin accessibility, and 3D genome structure are needed (Calzari *et al*., 2024; Cheong *et al*., 2023), along with single-cell epigenomics to uncover cell-type-specific alterations driving persistent symptoms (Nikesjö *et al*., 2025).

Most transcriptomic studies use bulk blood RNA sequencing, which can obscure signals from specific cell populations (Blankestijn *et al*., 2024; Zhang *et al*., 2023). Based on our review, more single-cell transcriptomics datasets should be created to resolve how individual cell types are reprogrammed in Long COVID (Gerayeli *et al*., 2024; Vaivode *et al*., 2024), and longitudinal profiling is needed to distinguish transient gene expression changes from persistent dysfunction (Phetsouphanh *et al*., 2024; Gómez-Carballa *et al*., 2025).

Proteomic analyses face technical limitations, including partial proteome coverage (Kruger *et al*., 2022; Medori *et al*., 2023). We recommend that future efforts should incorporate unbiased, high-throughput mass spectrometry to discover novel biomarkers (Klein *et al*., 2023; Suski *et al*., 2024) and improve quantification methods from relative to absolute protein quantification (Mohammed *et al*., 2024).

Metabolomic studies have identified disturbances in Long COVID patients but are often confounded by factors like diet, microbiome, and medication use (Ansone *et al*., 2024; López-Hernández *et al*., 2023). In our assessment, future research should aim for broader metabolite coverage using both targeted and untargeted approaches (Cysique *et al*., 2023; Machado *et al*., 2024; Martínez *et al*., 2024), with more rigorous study designs and absolute quantification methods.

A holistic understanding of Long COVID will appear from multi-omics integration. Each data type provides different perspectives of the same problem, with genomics identifying predispositions, transcriptomics and proteomics revealing active pathways, and metabolomics reflecting downstream biochemical effects (Ai *et al*., 2024; Lin *et al*., 2024). Our findings suggest that advanced algorithms should be applied to combine these diverse data types from the same patients (Wang *et al*., 2023), with standardized pipelines and network-based methods to visualize and interpret the combined datasets (Chen *et al*., 2024).

Future Long COVID research must integrate molecular profiling with wider data sources. Clinical data and advanced imaging modalities should be systematically collected alongside omics data from the same patients (Munblit *et al*., 2021; Thompson *et al*., 2022), allowing researchers to correlate molecular findings with clinical trajectories. From our analysis, incorporating pulmonary function tests, neurocognitive assessments, and wearable device data will better understand Long COVID (Holdsworth *et al*., 2022; Mancini *et al*., 2021). Standardizing data collection timing is crucial, as variability can lead to inconsistent results (Peppercorn *et al*., 2023; Saito *et al*., 2024; Taenzer *et al*., 2023). Longitudinal cohorts with regular follow-ups are essential to build models relating molecular dysfunction to organ system changes and patient-reported outcomes.

Analytical innovation is key to extracting actionable knowledge from complex Long COVID data. We believe causal inference techniques like MR can help distinguish correlation from causation (Pinero *et al*., 2025). Future applications should include Bayesian networks and Granger causality analyses to untangle complex cause-effect relationships and prioritize causal biomarkers and pathways.

Deep learning techniques can detect patterns in high-dimensional datasets, with unsupervised models discovering latent patient clusters (Liew *et al*., 2024). Autoencoder-based integration has identified distinct Long COVID patient endotypes (Ai *et al*., 2024; Klein *et al*., 2023). Based on our review, future research should emphasize explainable AI and ensure model generalizability while addressing sample size limitations and class imbalance.

Additional analytical techniques include network analysis to understand how dysregulated molecules interconnect (Li *et al*., 2025a; Lv *et al*., 2022), unsupervised clustering to discover patient subgroups (Ren *et al*., 2024; Su *et al*., 2022), and temporal modeling to map recovery patterns (Phetsouphanh *et al*., 2024).

AI agents, particularly Large Language Models (LLM), show high potential in healthcare applications. These systems can extract insights from scientific literature, assist in drug discovery, and reason over multi-modal data. The Virtual Lab provides an example of AI-driven research collaboration for designing nanobody binders against SARS-CoV-2 variants (Sawnson *et al*., 2024). In our view, this approach could be extended to biomarker discovery and personalized treatment strategies for Long COVID.

Computational analyses must be followed by experimental validation. Our analysis shows that in vitro studies using patient-derived immune cells can verify predicted molecular effects (Baillie *et al*., 2024; Kruger *et al*., 2022), while organoid models can investigate tissue-specific hypotheses (Kusakabe *et al*., 2023). Animal models allow testing of computationally identified pathways, and candidate drug compounds from in silico screens should be evaluated in appropriate models (Becker *et al*., 2020; Gupta *et al*., 2024). Validation across independent cohorts and with orthogonal assays is critical for establishing robust biomarkers.

The final goal is to apply computational insights toward precision medicine. We observe that stratifying patients into distinct endotypes based on molecular and clinical profiles has already begun (Liew *et al*., 2024; Woodruff *et al*., 2023), with studies identifying subsets of patients showing predominant immune activation, autonomic dysregulation, or metabolic imbalance (An *et al*., 2023; Cervia-Hasler *et al*., 2024). These endotypes could enable personalized management strategies (Dani *et al*., 2020).

Predictive modeling could identify individuals at risk of developing Long COVID after acute infection using genetic risk scores, acute infection biomarkers, and comorbidity data. ML algorithms integrating multi-omic data have been proposed as clinical decision-support tools (Su *et al*., 2022; Yong *et al*., 2023). From our perspective, the development of diagnostic assays based on confirmed biomarkers could objectively diagnose Long COVID and stratify patients (Gu *et al*., 2023; Klein *et al*., 2023).

Computational findings are also pointing to therapeutic targets, with network analyses highlighting dysregulated pathways like complement activation, inflammation, and clotting that might be addressed through drug repurposing (Baillie *et al*., 2024; Woodruff *et al*., 2023; Gupta *et al*., 2024). Our review suggests that adaptive clinical trial designs could evaluate multiple therapies in parallel, guided by biomarkers. The feedback from these trials will refine computational models, creating a cycle where treatment responses inform better predictions of what works for whom.

In the long term, we envision that clinicians will be able to classify Long COVID patients via omics tests and clinical data and select targeted interventions informed by that profile. This vision requires interdisciplinary collaboration between computational scientists, experimental biologists, and clinicians but promises significantly improved outcomes for those suffering from Long COVID.

By addressing current limitations and working on future directions—from multi-omics and big-data analytics to rigorous validation and translation—long COVID studies can generate robust, actionable insights for more reliable biomarkers, mechanistic understanding, and personalized therapies. In our assessment, integrating comprehensive data sources with cutting-edge computational methods, followed by careful experimental and clinical validation, will ensure that discoveries truly benefit patients, moving us closer to improving Long COVID symptoms through evidence-driven care and targeted interventions.

## 5. Conclusion

This study reviewed the computational methods for identifying biomarkers, predicting outcomes, and finding potential treatments for Long COVID. Our analysis covered multiple omics approaches, from genomic studies identifying the FOXP4 locus (Lammi *et al*., 2023) to proteomic analyses revealing 162-190 differentially expressed proteins (Peppercorn *et al*., 2023; Woodruff *et al*., 2023). The review highlighted successful prediction methods, including ML models achieving high accuracy in distinguishing Long COVID subtypes (Cervia-Hasler *et al*., 2024; Woodruff *et al*., 2023) and identifying critical biomarkers such as complement proteins and vascular transformation markers (Baillie *et al*., 2024; Maitray A Patel *et al*., 2022).

We examined various computational approaches for treatment discovery, including drug repurposing strategies that identified FDA-approved drugs targeting specific genetic markers (García-Hidalgo *et al*., 2022) and pathway analyses revealing potential therapeutic targets in the complement and inflammatory cascades (Cervia-Hasler *et al*., 2024; Yin *et al*., 2024). We also emphasized the importance of patient stratification, with studies identifying distinct endotypes that could guide personalized treatment approaches (An *et al*., 2023; Yong & Liu, 2022).

However, our analysis revealed significant limitations in current research, including small sample sizes, varying case definitions, and limited longitudinal data. Future studies should focus on larger, more diverse cohorts, standardized definitions, and extended follow-up periods. Integrating multi-omics data and advanced computational methods will be crucial for developing more effective diagnostic and therapeutic strategies.

By providing a comprehensive overview of computational methods, data resources, and current findings, this review aims to guide future research in Long COVID. We aim this work will support the development of more targeted interventions and ultimately improve outcomes for Long COVID patients.

## Data Availability

All data produced in the present work are contained in the manuscript.

https://docs.google.com/document/d/10o9zwSUeIC7JRu7bCF1SjlBeguNOlQo3/edit?usp=sharing&ouid=109216076945524164005&rtpof=true&sd=true

## Acknowledgements

This work was supported by STEM, University of South Australia (UniSA).

## Glossary

Long COVID: Persistent symptoms extending beyond 4 weeks from initial SARS-CoV-2 infection.
PASC: Post-Acute Sequelae of COVID-19, an alternative term for Long COVID.
Omics: High-throughput molecular data types.
GWAS: Genome-Wide Association Study, analysis of genetic variants across genomes.
EWAS: Epigenome-Wide Association Study, analysis of epigenetic modifications.
DMR: Differentially Methylated Region, area of DNA with altered methylation patterns.
scRNA-seq: Single-cell RNA sequencing, a method to analyze gene expression in individual cells.
DEG: Differentially Expressed Gene, gene with significantly altered expression levels.
DEP: Differentially Expressed Protein, protein with significantly altered abundance.
Multi-omics: Integration of multiple molecular data types for comprehensive analysis.
Endotype: Biological subtype of a condition defined by distinct molecular mechanisms.

