## Supplementary material for "Omics-Based Computational Approaches for Biomarker Identification, Prediction, and Treatment of Long COVID": SM1

### Supplementary Materials

#### 1. Supplementary Table S1: Search Methodology

##### Databases

This review was conducted using the following databases (accessed up to June 2, 2024):

- Google Scholar (https://scholar.google.com)
- PubMed (https://pubmed.ncbi.nlm.nih.gov)
- Gene Expression Omnibus (GEO) (https://www.ncbi.nlm.nih.gov/geo)
- European Nucleotide Archive (ENA) (https://www.ebi.ac.uk/ena)
- Sequence Read Archive (SRA) (https://www.ncbi.nlm.nih.gov/sra)
- COVID-19 Data Portal (https://www.COVID19dataportal.org)

##### Search Terms

We employed a comprehensive set of search terms organized by -omics domain to capture the full spectrum of molecular research on Long COVID. The complete search strings used for each -omics domain is detailed below:

**Genomics**

("genomics" OR "whole genome sequencing" OR "genome-wide association study" OR "GWAS" OR "genetic variants" OR "SNP analysis" OR "epigenomics" OR "genetic predisposition" OR "genetic risk factors" OR "functional genomics" OR "gene-environment interaction" OR "pharmacogenomics" OR "time-series" OR "tissues" OR "epigenetic modifications" OR "drug response" OR "environmental response" OR "functional validation") AND ("long-COVID" OR "long haul COVID" OR "long-haulier" OR "post-acute sequelae of SARS-CoV-2 infection" OR "PASC" OR "post-COVID syndrome" OR "chronic COVID syndrome" OR "long-term COVID effects" OR "COVID aftermath" OR "post-COVID complications")

**Epigenomics**

("epigenomics" OR "DNA methylation" OR "histone modification" OR "chromatin remodeling" OR "non-coding RNA" OR "miRNA expression" OR "epigenetic regulation" OR "epigenetic profiling" OR "epigenome-wide association study" OR "EWAS" OR "epigenetic markers" OR "epigenetic changes" OR "time-series" OR "tissues" OR "epigenetic response" OR "drug response" OR "environmental response" OR "functional validation") AND ("long-COVID" OR "long haul COVID" OR "long-haulier" OR "post-acute sequelae of SARS-CoV-2 infection" OR "PASC" OR "post-COVID syndrome" OR "chronic COVID syndrome" OR "long-term COVID effects" OR "COVID aftermath" OR "post-COVID complications")

**Transcriptomics**

("gene expression" OR "microarray" OR "RNA-seq" OR "bulk RNA sequencing" OR "single-cell RNA sequencing" OR "single-cell" OR "time-series" OR "tissues" OR "peripheral blood mononuclear cells" OR "nasopharyngeal swabs" OR "lung tissue" OR "functional genomics" OR "epigenetic response" OR "drug response" OR "environmental response" OR "functional validation") AND ("long-COVID" OR "long haul COVID" OR "long-haulier" OR "post-acute sequelae of SARS-CoV-2 infection" OR "PASC" OR "post-COVID syndrome" OR "chronic COVID syndrome" OR "long-term COVID effects" OR "COVID aftermath" OR "post-COVID complications" OR "post-viral fatigue")

**Proteomics**

("proteomics" OR "mass spectrometry" OR "protein expression" OR "protein profiling" OR "phosphoproteomics" OR "glycoproteomics" OR "metaproteomics" OR "quantitative proteomics" OR "shotgun proteomics" OR "functional proteomics" OR "label-free proteomics" OR "targeted proteomics" OR "time-series" OR "tissues" OR "post-translational modifications" OR "drug response" OR "environmental response" OR "functional validation") AND ("long-COVID" OR "long haul COVID" OR "long-haulier" OR "post-acute sequelae of SARS-CoV-2 infection" OR "PASC" OR "post-COVID syndrome" OR "chronic COVID syndrome" OR "long-term COVID effects" OR "COVID aftermath" OR "post-COVID complications")

**Metabolomics**

("metabolomics" OR "metabolic profiling" OR "mass spectrometry" OR "nuclear magnetic resonance spectroscopy" OR "NMR spectroscopy" OR "liquid chromatography-mass spectrometry" OR "LC-MS" OR "gas chromatography-mass spectrometry" OR "GC-MS" OR "metabolite analysis" OR "lipidomics" OR "metabonomics" OR "metabolic response" OR "time-series" OR "tissues" OR "drug response" OR "environmental response" OR "functional validation") AND ("long-COVID" OR "long haul COVID" OR "long-haulier" OR "post-acute sequelae of SARS-CoV-2 infection" OR "PASC" OR "post-COVID syndrome" OR "chronic COVID syndrome" OR "long-term COVID effects" OR "COVID aftermath" OR "post-COVID complications" OR "post-viral fatigue")

**Multi-omics**

("multi-omics" OR "integrative omics" OR "systems biology" OR "multi-layer omics" OR "transcriptomics" OR "genomics" OR "epigenomics" OR "proteomics" OR "metabolomics" OR "lipidomics" OR "metagenomics" OR "multi-modal data integration" OR "cross-omics analysis" OR "multi-omics profiling" OR "functional multi-omics" OR "time-series" OR "tissues" OR "drug response" OR "environmental response" OR "functional validation") AND ("long-COVID" OR "long haul COVID" OR "long-haulier" OR "post-acute sequelae of SARS-CoV-2 infection" OR "PASC" OR "post-COVID syndrome" OR "chronic COVID syndrome" OR "long-term COVID effects" OR "COVID aftermath" OR "post-COVID complications" OR "post-viral fatigue")

#### Search Strategy

Each search string was applied to all databases to ensure comprehensive coverage. Additional filters were applied for specialized repositories (GEO, ENA, SRA) to identify datasets specifically related to Long COVID studies. The COVID-19 Data Portal was searched using simplified versions of these terms to accommodate its specific search functionality.

#### Inclusion and Exclusion Criteria

Studies were included if they:

- Employed at least one -omics approach.
- Focused on Long COVID or post-acute sequelae of SARS-CoV-2 infection.
- Were published in English.
- Had available full-text or complete dataset information.

Studies were excluded if they:

- Focused solely on acute COVID-19 without long-term follow-up.
- Were review articles without original data.
- Used only clinical parameters without molecular analyses.
- Had significant methodological limitations.

#### 2. Supplementary Table S2: Data Sources and Computational Methods in Long COVID Research

**Note: Refer to SM2.xlsx for access to the entire table.**

This supplementary table provides a comprehensive catalog of datasets and computational approaches used in Long COVID research as of March 2025. The table includes detailed information on:

- Study characteristics (author, year, and cohort size)
- Sample types and collection time points
- Omics data types (genomics, epigenomics, transcriptomics, proteomics, metabolomics, and multi-omics)
- Specific assay methodologies and platforms
- Computational analysis pipelines and software tools
- Statistical approaches and correction methods
- Key biological findings and identified biomarkers
- Integration methods for multi-omics studies

This resource is designed to facilitate data discovery and meta-analysis for researchers investigating the molecular mechanisms of Long COVID. The table will be continuously updated as new studies emerge in this rapidly evolving field.
